# Genome-wide investigation of maximum habitual alcohol intake (MaxAlc) in 247,755 European and African Ancestry U.S. Veterans informs the relationship between habitual alcohol consumption and alcohol use disorder

**DOI:** 10.1101/2022.05.02.22274580

**Authors:** J. D. Deak, D. F. Levey, F. R. Wendt, H. Zhou, M. Galimberti, H. R. Kranzler, J. M. Gaziano, M.B. Stein, R. Polimanti, The Million Veteran Program, J. Gelernter

## Abstract

**Importance:** Alcohol genome-wide association studies (GWAS) have generally focused on alcohol consumption and alcohol use disorder (AUD); few have examined habitual drinking behaviors like maximum habitual alcohol intake (MaxAlc).

**Objective:** Identify MaxAlc loci and elucidate the genetic architecture across alcohol traits.

**Design:** The MaxAlc GWAS was performed in Million Veteran Program (MVP) participants enrolled from January 10, 2011 to September 30, 2020. Ancestry-specific GWAS were conducted in European (EUR) (n=218,623) and African (AFR) (n=29,132) ancestry subjects, then meta-analyzed (N=247,755). Linkage-disequilibrium score regression (LDSC) was used to estimate SNP-heritability and genetic correlations (r_g_) with other alcohol and psychiatric traits. Genomic structural equation modeling (gSEM) was used to evaluate genetic relationships between MaxAlc and other alcohol traits. Mendelian randomization (MR) was used to examine causal relationships. MTAG (multi-trait analysis of GWAS) was used to analyze MaxAlc and problematic alcohol use (PAU) jointly.

**Setting:** The study was performed in a sample of U.S military Veterans.

**Participants:** Participants were 92.68% male and had mean age=65.92 (*SD*=11.70). 36.92% reported MaxAlc ≥ the binge-drinking threshold.

**Main Outcomes(s) and Measure(s):** MaxAlc was defined from survey item: “in a typical month, what is/was the largest number of drinks of alcohol you may have had in one day?” with ordinal responses from 0 ≥ 15 drinks.

**Results:** The MaxAlc GWAS resulted in 15 genome-wide significant (GWS) loci. Top associations in EUR and AFR were with known functional variants *ADH1B*\*rs1229984 (*p*=3.12×10^−104^) and rs2066702 (*p*=6.30×10^−17^), respectively. Multiple novel associations were found. The SNP-heritability was 6.65% (s.e.=0.41%) in EUR and 3.42% (s.e.=1.46%) in AFR. MaxAlc was positively correlated with PAU (r_g_=0.79; *p*=3.95×10^−149^) and AUD (r_g_=0.76; *p*=1.26×10^−127^), and had negative r_g_ with “alcohol usually taken with meals” (r_g_=-0.53; *p*=1.40×10^−50^). For psychiatric traits, MaxAlc had the strongest r_g_ with suicide attempt (r_g_=0.40; *p*=3.02×10^−21^). gSEM supported a two-factor model with MaxAlc loading on a factor with PAU and AUD, and other alcohol consumption measures loading a separate factor. MR supported a small causal effect of MaxAlc on the liver enzyme gamma-glutamyltransferase (β=0.012; *p*=2.66×10^−10^). MaxAlc MTAG resulted in 31 GWS loci.

**Conclusions and Relevance:** MaxAlc closely aligns genetically with the etiology of problematic alcohol use traits.

**Key Points:** *Question:* What is the genetic etiology of maximum habitual alcohol intake (MaxAlc) and how does it compare to other alcohol consumption measures.

*Findings:* This MaxAlc study in 247,455 European and African ancestry individuals identified 15 genome-wide significant loci, including multiple novel associations. MaxAlc was strongly genetically correlated (r_g_) with measures of alcohol-related problems, demonstrated significantly different r_g_ with psychiatric traits compared to other alcohol consumption traits, and loaded on a factor with alcohol problem traits while alcohol consumption state measures loaded on a separate factor.

*Meaning:* MaxAlc is genetically different from trait consumption measures in relation to problematic alcohol use.

## Introduction

Progress has been made in understanding the genetic etiology of many alcohol-related traits(1-2) including alcohol use disorder (AUD)(3-4), forms of problematic alcohol use (PAU)(4-5), and measures of alcohol consumption(3,5-6). A major contribution that genome-wide association studies (GWAS) have made to understanding the genetics of alcohol traits is the finding that the genetic architecture of AUD differs from alcohol use measures(2). Genetic correlation analyses have overturned the idea that alcohol use traits differ from AUD only in degree of use by showing consistently that they differ in nature too(3-5). Quantity-frequency measures like the AUDIT-C (Alcohol Use Disorders Identification Test–Consumption score) correlate genetically with positive metabolic outcomes but not psychiatric disorders, while AUD correlates genetically with psychiatric disorders(3-4). Although these findings have been replicated, they are not well understood and warrant further study(7). Few well-powered efforts have examined the genetic influences across levels of habitual alcohol consumption—an important area of investigation given that higher levels of alcohol intake often lead to increased negative alcohol outcomes.

We aim to improve understanding of the genetic etiology of habitual alcohol consumption by greatly increasing the sample size of GWAS of maximum habitual alcohol intake (MaxAlc)(8) in European (EUR) and African (AFR) ancestry participants in the Million Veteran Program (MVP), and examining the similarities and differences between MaxAlc and other alcohol traits. MaxAlc can be distinguished from similar phenotypes that measure the maximum number of drinks consumed in a lifetime drinking episode (“lifetime max-drinks”), in that MaxAlc captures habitual maximum alcohol intake (i.e., “in a typical month”), while “lifetime max-drinks” may capture consumption on only a single occasion, often being influenced by “milestone” drinking events (e.g., 21^st^ birthday celebrations)(9). Thus, because MaxAlc is more representative of routine behaviors, it is likely more clinically meaningful and representative of behavior that could result in more negative alcohol-related outcomes than occasional binge-drinking or lower levels of alcohol consumption(10).

The current study is the largest GWAS of MaxAlc to date. The larger sample enables us to estimate the heritability of MaxAlc and examine its genetic correlations with traits from large studies of alcohol consumption and alcohol-related problems. We used genomic structural equation modeling (gSEM) to evaluate the overarching genetic relations between MaxAlc and other alcohol traits. To investigate the impact of MaxAlc on long-term alcohol-related disease, we conducted Mendelian randomization (MR) to examine causal relationships between MaxAlc and liver enzyme levels. Together, these analyses provide a better understanding of the genetic basis of regular heavy drinking and how it compares to the genetics of other alcohol consumption measures and AUD.

## Method

### Participants and Phenotyping

MVP participants of EUR and AFR ancestry with MaxAlc phenotypic and genotypic data (EUR: n=218,623; AFR: n=29,132) were included resulting in a total sample of 247,755 subjects. MaxAlc was defined using information from the MVP Lifestyle Survey item: “in a typical month, what is/was the largest number of drinks of alcohol you may have had in one day?” with ordinal response options ranging from 0 to ≥ 15 drinks (**Supplemental Figure1**). We removed ≥ second-degree relatives pairwise based on a kinship coefficient >0.0884, prioritizing the retention of the individual with the higher reported MaxAlc value (the heavier drinking member of the pair of related individuals). The total sample was 92.68% male, had a mean age of 65.92 (*SD*=11.70), and had 36.92% who reported MaxAlc > the binge-drinking threshold(11) (females: ≥4 drinks; males ≥5 drinks)(**Supplemental Table1**; **Supplemental Figure2**). The study was approved by the Central VA institutional review board (IRB) and site-specific IRBs, including Yale University School of Medicine and VA Connecticut, and was conducted in accordance with all relevant ethical regulations.

### Genotyping, Imputation, and Quality Control

Genotyping and imputation for MVP are described in **supplemental materials**.

### Ancestry-specific GWAS and Cross-ancestry GWAS meta-analysis

Individual GWAS were conducted on the ordinal trait in the respective MVP samples of EUR and AFR ancestry subjects using a linear regression model implemented in PLINK 2.0(12). Age, sex, and the first 10 within-ancestry genetic PCs were included as covariates. Ancestry-specific GWAS were then meta-analyzed across ancestries using inverse-variance weighting in METAL(13). 1KG Phase 3(14) was used as a reference panel to determine EUR and AFR linkage disequilibrium (LD) structure. Independent GWAS loci were identified using a linkage-disequilibrium (LD) threshold of r^2^=0.1 to determine lead SNPs at each genome-wide significant (GWS) locus(15). Variants were assigned to the nearest gene based upon physical position (<10 kb from the assigned gene). Additional gene-mapping techniques using eQTL (expression quantitative trait locus) and 3D chromatin interactions [Hi-C]) data are described in **supplemental materials** (**Supplement Figures18-19**).

### Gene, Gene-set, and Tissue-Specific Gene Expression Analysis

Gene, gene-set, and gene expression analyses are described in **supplemental materials**.

### SNP-heritability and genetic correlations

LD score regression (LDSC)(16) was used to estimate SNP-heritability (h^2^_SNP_) and to characterize genetic correlations (r_g_) with a set of alcohol, psychiatric, and medical traits. Genetic correlations were examined for MaxAlc with alcohol use disorder (AUD)(3), problematic alcohol use (PAU)(4), drinks per week (DPW)(6), a UK Biobank (UKB) measure of “when you drink alcohol is it usually with meals?” (UKB Field ID 1618), and the Alcohol Use Disorders Identification Test (AUDIT) total (AUDIT-T), problems (AUDIT-P), and consumption (AUDIT-C) scales(3,5) in EUR. Measures of AUDIT-C from two samples that have demonstrated differences in substance use problems and demographic composition, MVP and UKB, were included to examine distinctions between the measure across these populations. COV-LDSC(17) was used to estimate MaxAlc h^2^_SNP_ in AFR.

Large-scale GWAS of AUD and PAU, unlike alcohol consumption measures, have shown positive genetic correlations with psychiatric disorders(3-4). GWAS have not yet been employed to examine patterns of genetic correlation between psychopathology and heavier alcohol consumption (e.g., MaxAlc) in comparison to AUD and other consumption measures. Thus, we performed genetic correlation analyses between the included alcohol traits and GWAS of anxiety(18), depression(19), PTSD(20), schizophrenia(21), and suicide attempt(22).

### Genomic Structural Equation Modeling of alcohol traits

Genomic-SEM (gSEM)(23) was used to evaluate the shared polygenic architecture of MaxAlc, alcohol use problems, and other alcohol consumption measures using GWAS summary statistics. Exploratory factor analysis (EFA) was used to evaluate one-, two-, and three-factor model fit. Confirmatory factor analysis (CFA) was then performed to evaluate model convergence and factor loadings using conventional model fit indices(23). CFA models were estimated using diagonally weighted least squares estimation and a smoothed genetic covariance matrix to guard against non-positive definite scenarios. The 1kGP Phase 3 EUR reference panel was used to calculate LD(14).

### Mendelian Randomization (MR) Analyses

We used Mendelian randomization (MR) to examine causal relationships between MaxAlc and alcohol-related liver enzyme levels with available GWAS data (ALP [alkaline phosphatase], ALT [alanine transaminase], and GGT [gamma-glutamyl transferase])(24) using inverse-variance weighted (IVW), weighted median, and MR-Egger methods in TwoSampleMR(25) and MR-PRESSO(26). MR analysis was limited to liver enzymes demonstrating a significant genetic correlation with MaxAlc. To ensure sufficient genetic instruments to test for causal relationships, we included LD-independent MaxAlc loci reaching a suggestive significance threshold of *p*<5.0×10^−5^ (nSNPs=171). Multiple methods were used to evaluate method-specific biases to determine consistent effect estimates. Sensitivity analyses, leave-one-out analyses, and outlier removal were all evaluated. The MR-Egger intercept test was used to evaluate the presence of horizontal pleiotropy. MR-PRESSO(26) was used to identify genetic instrument outliers that show horizontal pleiotropy for removal. MR-RAPS (MR-Robust Adjusted Profile Score)(27) was used to test for biased effect estimates due to the inclusion of “suggestive significance” SNPs as genetic instruments.

### Multi-trait analysis of MaxAlc and PAU

A joint analysis of EUR MaxAlc and PAU(4) GWAS summary statistics was performed in MTAG (Multi-trait analysis of GWAS)(28). The MTAG analysis is described in **supplemental materials**.

## Results

### Ancestry-specific GWAS and Cross-ancestry GWAS meta-analysis

The MaxAlc GWAS resulted in 15 LD-independent GWS (*p*≤5.0×10^−08^) loci across the respective analyses (**Figure1**; **Table1**). There were 10 GWS loci in the EUR analysis, 2 GWS loci in the AFR-specific analysis, and 3 additional GWS loci in the cross-ancestry meta-analysis that did not reach GWS in the ancestry-specific analyses.

**Table 1.** **GWS MaxAlc loci in the (A) cross-ancestry GWAS; (B) EUR GWAS, (C) AFR GWAS, and (D) MaxAlc MTAG GWAS**.

**A. Cross-ancestry MaxAlc GWS ( $p \leq 5.00E-08$ ) lead SNPs**
| Chr | Position | MarkerName | A1 | A2 | Nearest Gene | Direction | P-value | nSNPs | nGWASSNPs | nIndSigSNPs | nLeadSNPs |
| --- | --- | --- | --- | --- | --- | --- | --- | --- | --- | --- | --- |
| 4 | 100239319 | rs1229984 | C | T | *ADH1B* | -? | 2.46E-101 | 529 | 393 | 22 | 9 |
| 4 | 103188709 | rs13107325 | T | C | *SLC39A8* | -- | 9.40E-15 | 23 | 23 | 4 | 1 |
| 17 | 44305199 | rs570285046 | C | T | *CRHR1* | ++ | 2.12E-14 | 3590 | 3538 | 29 | 3 |
| 7 | 135162897 | rs2551763 | T | C | *CNOT4* | -? | 1.29E-09 | 29 | 18 | 1 | 1 |
| 9 | 30359634 | rs13297433 | A | G | *snoU13* | ++ | 2.01E-09 | 76 | 73 | 2 | 1 |
| 11 | 47392114 | rs7928419 | A | G | *SPI1* | ++ | 2.29E-09 | 196 | 172 | 3 | 1 |
| 1 | 7830262 | rs2153733 | C | T | *CAMTA1* | +? | 1.36E-08 | 1 | 1 | 1 | 1 |
| 19 | 37504953 | rs826323 | A | G | *ZNF420* | ++ | 1.39E-08 | 162 | 153 | 1 | 1 |
| 9 | 33094361 | rs56175925 | A | G | *B4GALT1* | ?+ | 1.82E-08 | 5 | 5 | 1 | 1 |
| 10 | 10704659 | rs34165302 | C | CA | *RP1-186E20.1* | -? | 2.08E-08 | 87 | 35 | 1 | 1 |
| 10 | 110572259 | rs1577857 | G | T | *RP11-655H13.2* | +? | 2.47E-08 | 100 | 82 | 1 | 1 |
| 2 | 104056769 | rs6752675 | A | G | *AC092568.1* | -- | 2.57E-08 | 196 | 170 | 1 | 1 |
| 2 | 27730940 | rs1260326 | C | T | *GCKR* | -- | 2.63E-08 | 9 | 9 | 1 | 1 |
| 11 | 113392994 | rs2514218 | T | C | *DRD2* | -- | 4.59E-08 | 25 | 23 | 1 | 1 |

**B. EUR MaxAlc GWS ( $p \leq 5.00E-08$ ) Lead SNPs**
| Chr | Position | MarkerName | A1 | A2 | Nearest Gene | MAF | P-value | nSNPs | nGWASSNPs | nIndSigSNPs | nLeadSNPs |
| --- | --- | --- | --- | --- | --- | --- | --- | --- | --- | --- | --- |
| 4 | 100239319 | rs1229984 | C | T | *ADH1B* | 0.04 | 3.12E-101 | 420 | 275 | 14 | 8 |
| 4 | 103188709 | rs13107325 | T | C | *SLC39A8* | 0.08 | 6.78E-15 | 24 | 24 | 2 | 1 |
| 17 | 43810896 | rs77804065 | T | C | *CRHR1* | 0.23 | 6.04E-14 | 3839 | 3533 | 4 | 2 |
| 11 | 47676170 | rs7107356 | G | A | *AGBL2* | 0.5 | 1.43E-10 | 422 | 313 | 3 | 1 |
| 7 | 135162897 | rs2551763 | T | C | *CNOT4* | 0.41 | 1.30E-09 | 43 | 19 | 1 | 1 |
| 1 | 7830262 | rs2153733 | C | T | *CAMTA1* | 0.19 | 1.36E-08 | 35 | 31 | 1 | 1 |
| 10 | 10714452 | rs145961009 | C | CTT | *RP1-186E20.1* | 0.22 | 1.73E-08 | 133 | 42 | 1 | 1 |
| 9 | 30346448 | rs4879424 | C | T | *snoU13* | 0.15 | 2.18E-08 | 190 | 169 | 1 | 1 |
| 10 | 110572259 | rs1577857 | G | T | *RP11-655H13.2* | 0.26 | 2.47E-08 | 122 | 102 | 1 | 1 |
| 2 | 27730940 | rs1260326 | C | T | *GCKR* | 0.42 | 4.53E-08 | 18 | 18 | 1 | 1 |

**C. AFR MaxAlc GWS ( $p \leq 5.00E-08$ ) Lead SNPs**
| Chr | Position | MarkerName | A1 | A2 | Nearest Gene | MAF | P-value | nSNPs | nGWASSNPs | nIndSigSNPs | nLeadSNPs |
| --- | --- | --- | --- | --- | --- | --- | --- | --- | --- | --- | --- |
| 4 | 100229017 | rs2066702 | A | G | *ADH1B* | 0.19 | 6.30E-17 | 109 | 103 | 7 | 1 |
| 9 | 33094361 | rs56175925 | A | G | *B4GALT1* | 0.02 | 1.83E-08 | 6 | 5 | 1 | 1 |

GENETIC INFLUENCES ON MAXIMUM HABITUAL ALCOHOL INTAKE D. MaxAlc MTAG GWS ( $p \leq 5.00E-08$ ) Lead SNPs
| Chr | Position | MarkerName | A1 | A2 | Nearest Gene | MAF | P-value | nSNPs | nGWASSNPs | nIndSigSNPs | nLeadSNPs |
| --- | --- | --- | --- | --- | --- | --- | --- | --- | --- | --- | --- |
| 4 | 100239319 | rs1229984 | T | C | *ADH1B* | 0.04 | 3.35E-176 | 1291 | 646 | 26 | 9 |
| 4 | 103188709 | rs13107325 | T | C | *SLC39A8* | 0.08 | 6.01E-36 | 75 | 26 | 6 | 1 |
| 2 | 27730940 | rs1260326 | T | C | *GCKR* | 0.41 | 2.72E-21 | 261 | 98 | 4 | 1 |
| 4 | 39423789 | rs4975013 | A | G | *KLB* | 0.39 | 1.44E-19 | 64 | 27 | 4 | 1 |
| 11 | 113364691 | rs17602038 | T | C | *DRD2* | 0.37 | 2.62E-16 | 70 | 36 | 3 | 2 |
| 11 | 47406592 | rs11039216 | T | C | *RP11-750H9.5* | 0.47 | 1.07E-15 | 519 | 245 | 5 | 1 |
| 17 | 44083402 | rs1991556 | A | G | *MAPT* | 0.23 | 2.25E-15 | 3854 | 2089 | 2 | 1 |
| 10 | 110572259 | rs1577857 | T | G | *RP11-655H13.2* | 0.26 | 6.44E-15 | 494 | 304 | 2 | 1 |
| 2 | 104361420 | rs9308868 | A | G | *AC013727.1* | 0.49 | 4.88E-13 | 518 | 329 | 3 | 1 |
| 14 | 58765903 | rs61974485 | T | C | *ARID4A* | 0.28 | 1.02E-12 | 117 | 5 | 1 | 1 |
| 2 | 45139904 | rs472140 | T | C | *RP11-89K21.1* | 0.42 | 1.11E-11 | 50 | 5 | 2 | 1 |
| 7 | 135119417 | rs2551774 | A | G | *CNOT4* | 0.41 | 1.50E-11 | 43 | 14 | 1 | 1 |
| 10 | 10688361 | rs7910201 | A | G | *RP1-186E20.1* | 0.25 | 6.36E-11 | 168 | 69 | 2 | 1 |
| 16 | 53834607 | rs7188250 | T | C | *FTO* | 0.41 | 8.06E-10 | 106 | 82 | 1 | 1 |
| 7 | 153487944 | rs2098112 | A | G | *DPP6* | 0.47 | 8.99E-10 | 112 | 2 | 1 | 1 |
| 19 | 49244219 | rs2307018 | A | C | *IZUMO1* | 0.43 | 9.98E-10 | 59 | 21 | 1 | 1 |
| 1 | 66503517 | rs1354063 | T | C | *PDE4B* | 0.47 | 1.21E-09 | 224 | 81 | 3 | 1 |
| 15 | 47681384 | rs8033799 | A | C | *CTD-2050N2.1* | 0.21 | 1.86E-09 | 157 | 65 | 2 | 1 |
| 5 | 153364650 | rs2245405 | A | G | *FAM114A2* | 0.45 | 2.24E-09 | 70 | 32 | 1 | 1 |
| 22 | 41789408 | rs4822028 | A | G | *TEF* | 0.23 | 7.97E-09 | 762 | 503 | 2 | 2 |
| 12 | 81602449 | rs1921035 | A | G | *ACSS3* | 0.48 | 8.44E-09 | 100 | 55 | 1 | 1 |
| 2 | 144225215 | rs13024996 | A | C | *RP11-570L15.2* | 0.36 | 8.56E-09 | 29 | 7 | 1 | 1 |
| 13 | 97019647 | rs1925104 | T | G | *HS6ST3* | 0.33 | 1.37E-08 | 100 | 56 | 1 | 1 |
| 8 | 21831385 | rs1484162 | A | G | *XPO7* | 0.41 | 1.49E-08 | 56 | 15 | 1 | 1 |
| 2 | 138197269 | rs10496771 | T | C | *THSD7B* | 0.25 | 1.56E-08 | 222 | 84 | 1 | 1 |
| 20 | 18546199 | rs6045466 | T | C | *LINC00493* | 0.47 | 1.92E-08 | 150 | 102 | 1 | 1 |
| 2 | 58042241 | rs1402398 | A | G | *CTD-2026C7.1* | 0.39 | 2.00E-08 | 268 | 55 | 2 | 1 |
| 7 | 117576675 | rs6466637 | A | G | *AC007568.1* | 0.28 | 2.00E-08 | 39 | 22 | 1 | 1 |
| 7 | 73015369 | rs13240065 | A | G | *MLXIPL* | 0.12 | 2.95E-08 | 66 | 46 | 1 | 1 |
| 2 | 204123832 | rs56059523 | T | C | *CYP20A1* | 0.11 | 3.17E-08 | 18 | 5 | 1 | 1 |
| 3 | 18793340 | rs9835977 | T | C | *AC144521.1* | 0.30 | 3.57E-08 | 79 | 44 | 1 | 1 |

**Figure 1.**
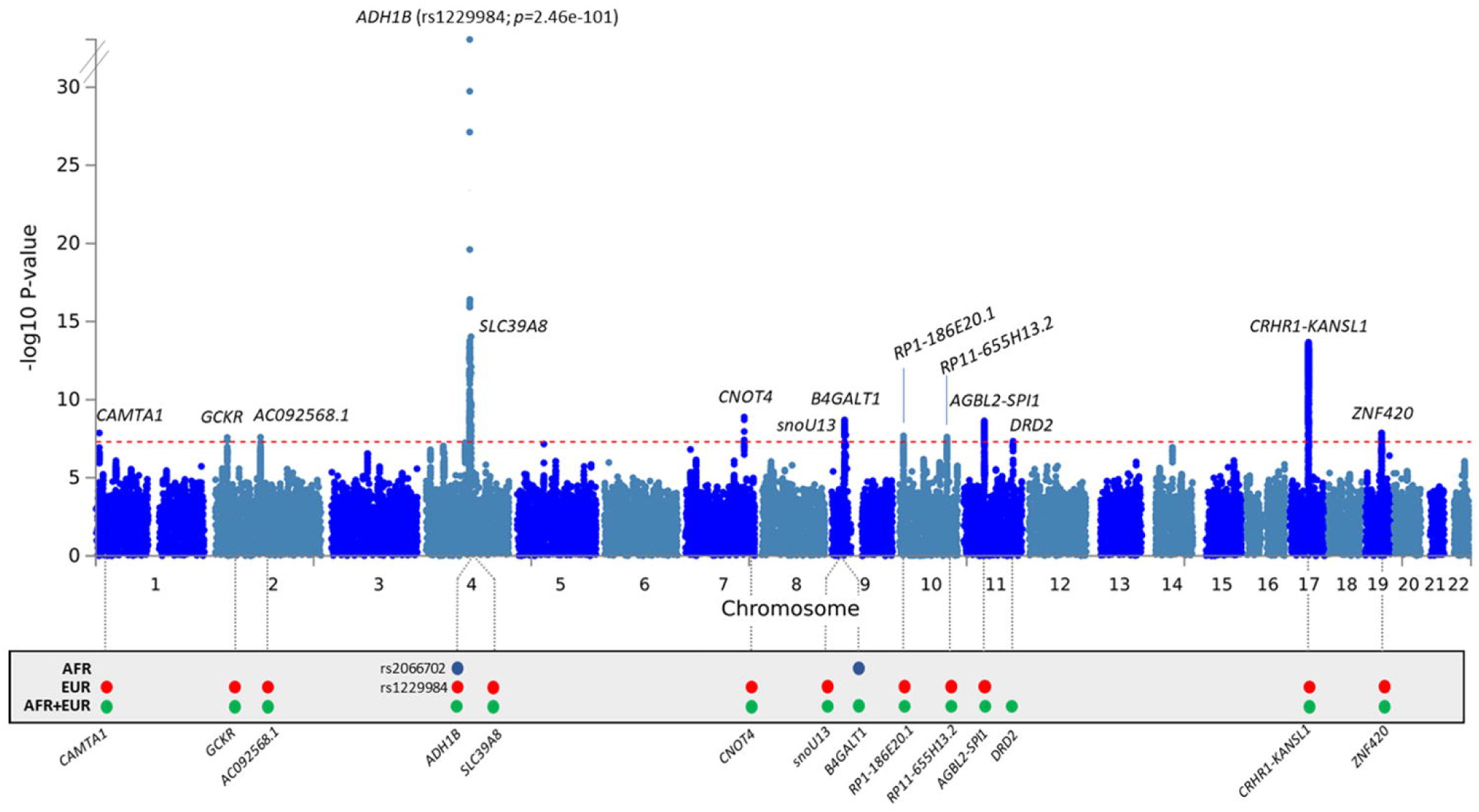
Manhattan plot of MaxAlc GWAS in 247,755 individuals of African (AFR) and European (EUR) ancestry. *Note:* Red-dashed line indicates genome-wide significance (GWS; 5.00×10^08^). Low panel: Blue circle indicates GWS in AFR; Red circle indicates GWS in EUR; Green circle indicates GWS in the cross-ancestry meta-analysis.

The top association in EUR was with the well-replicated functional *ADH1B* rs1229984 variant (*p*=3.12×10^−101^). Additional top GWS loci in EUR included *SLC39A8* (rs13107325; *p*=6.78×10^−15^), *CRHR1* (rs77804065; *p*=6.04×10^−14^), *AGBL2* (rs7107356; *p*=1.43×10^−10^), and *GCKR* (rs1260326; *p*=4.53×10^−08^). All loci in EUR remained GWS in the cross-ancestry meta-analysis, although there were differences in the lead SNPs identified in the two analyses (**Figure1; Table1**).

The top association in AFR was with the known AFR-specific functional locus at *ADH1B* (rs2066702; *p*=6.30×10^−17^). A second GWS association was found on chromosome 9 nearest *B4GALT1* (rs56175925; *p*=1.83×10^−08^)(**Figure1; Table1**).

In the cross-ancestry meta-analysis, three loci not identified in the ancestry-specific GWAS were GWS: rs826323, which maps to *ZNF420* (*p*=1.39×10^−08^) on chromosome 19; rs6752675 on chromosome 2 nearest *AC092568*.*1* (*p*=2.57×10^−08^); and *DRD2\**rs2514218 on chromosome 11 (*p*=4.59×10^−08^)(**Figure1; Table1**).

### Gene, Gene-set, and Tissue-Specific Gene Expression Analysis

Gene, gene-set, and gene expression results are reported in **supplemental materials**.

### SNP-heritability and genetic correlations

LDSC estimated an observed-scale MaxAlc h^2^_SNP_ of 6.65% (s.e.=0.41%) in EUR. LDSC estimates indicated that genome-wide inflation (λ_GC_=1.26) was likely indicative of the polygenic architecture of MaxAlc as supported by an LDSC intercept=0.99 (s.e.=0.004) and attenuation ratio <0. For AFR, COV-LDSC estimated h^2^_SNP_ =3.42% (s.e.=1.46%), a λ_GC_=1.03, LDSC intercept=0.99 (s.e.=0.007), and attenuation ratio <0.

In EUR, LDSC estimated a high r_g_ for MaxAlc with PAU (r_g_=0.79; *p*=3.95×10^−149^) and AUD (r_g_=0.76; *p*=1.26×10^−127^), a lower r_g_ with consumption traits (MVP AUDIT-C [r_g_=0.35; *p*=8.49×10^−15^]; DPW [r_g_=0.64; *p*=1.26×10^−103^]), and a negative r_g_ with “alcohol usually taken with meals” (r_g_ = −0.53; *p*=1.57×10^−52^)(**Figure2**). Among the psychiatric traits examined, MaxAlc had the highest r_g_ with suicide attempt (r_g_=0.40; *p*=3.02×10^−21^)(**Figure2; Supplemental Table14)**. The MaxAlc r_g_ with suicide attempt was significantly higher than the r_g_ between suicide attempt and all other consumption measures, but was not significantly different than the r_g_ between suicide attempt and AUD or PAU (**Supplemental Table14**). MaxAlc’s r_g_ with depression (r_g_=0.23; *p*=8.40×10^−16^), PTSD (r_g_=0.19; *p*=4.75×10^−05^), and anxiety (r_g_=0.15; *p*=5.60×10^−03^) were also significantly different from those observed for alcohol consumption traits. No significant differences between MaxAlc and other consumption measures were observed for schizophrenia. All alcohol trait correlations with the core psychiatric traits included in the current study are reported (**Supplemental Table14**).

**Figure 2.**
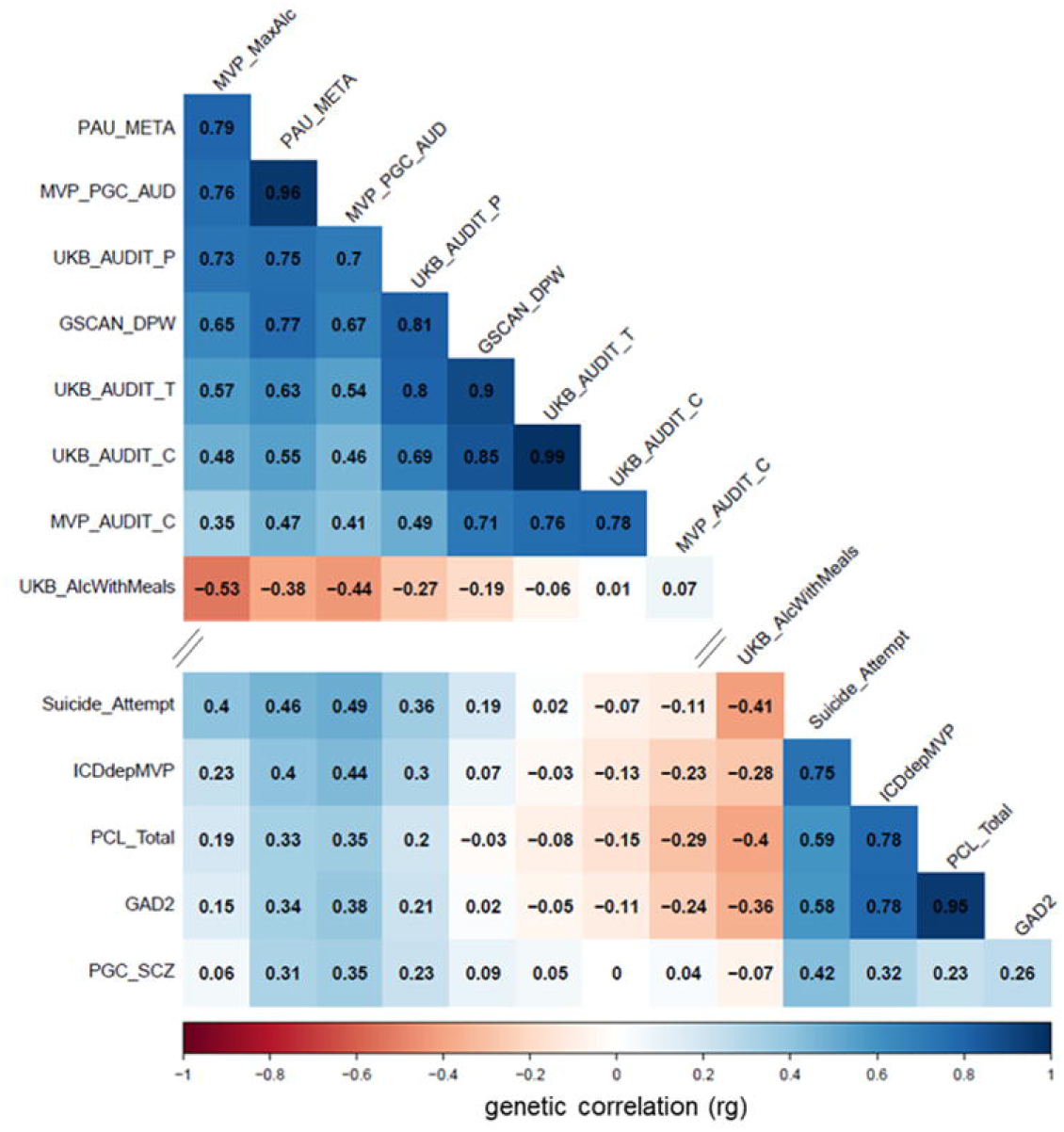
Genetic correlations between MVP MaxAlc, alcohol traits, and psychiatric outcomes. *Note:* Abbreviations (along diagonal): Maximum habitual alcohol intake (MaxAlc); Problematic alcohol use (PAU); Alcohol use disorder (AUD); Alcohol Use Disorders Identification Test-Problems (AUDIT-P); Drinks per week (DPW); AUDIT-Total (AUDIT-T); AUDIT-Consumption (AUDIT-C); ICD-based Depression (ICDdepMVP); PTSD Checklist -Total score (PCL_Total): Generalized Anxiety Disorder 2-item (GAD-2).

### Genomic Structural Equation Modeling

EFA and CFA were conducted using gSEM for MaxAlc and a range of previously-published alcohol traits ranging from AUD to alcohol consumption (AUDIT-C and DPW).

Using EFA to examine different models showed that a common factor model did not adequately capture the relationship across all alcohol traits (cumulative variance explained=0.59)(**Supplemental Table13**). The cumulative variance explained by the two-factor model was 0.82 (Factor 1=0.42; Factor 2=0.40), with high sum of squared (SS) loadings for both Factor 1 (Factor 1 SS=2.91) and Factor 2 (Factor 2 SS=2.83). The three-factor model explained less variance than the two-factor model (cumulative variance=0.80; Factor1=0.35, Factor 2=0.32, Factor 3=0.14) and had SS loadings of 2.42 (Factor 1), 2.23 (Factor 2), and 0.98 (Factor 3). Based on these findings, Factor 3 was not worth retaining given that the three-factor model accounted for less variance and the SS loading of Factor 3 was < 1.0. The two-factor model best fit the data and was evaluated further using CFA.

CFA converged on a two-factor model (correlation=0.67) and demonstrated strong fit across the included alcohol traits (CFI [Comparative Fit Index]=0.98; χ2= 238.38; AIC [Aikake Information Criterion]=270.38; SRMR [Standardized Root Mean Square Residual]=0.05) (**Figure3; Supplemental Table14**). MaxAlc loaded most strongly on Factor 1 (loading=0.85±0.05) along with PAU (loading=0.93±0.03) and AUD (loading=1.00±0.03). MVP AUDIT-C (loading=0.72±0.04), UKB AUDIT-C (loading=0.84±0.03), DPW (loading=1.02±0.03), and AUDIT-P (loading=0.46±0.06) loaded most strongly on Factor 2, though AUDIT-P also loaded equally well on Factor 1 (loading=0.46±0.05).

**Figure 3.**
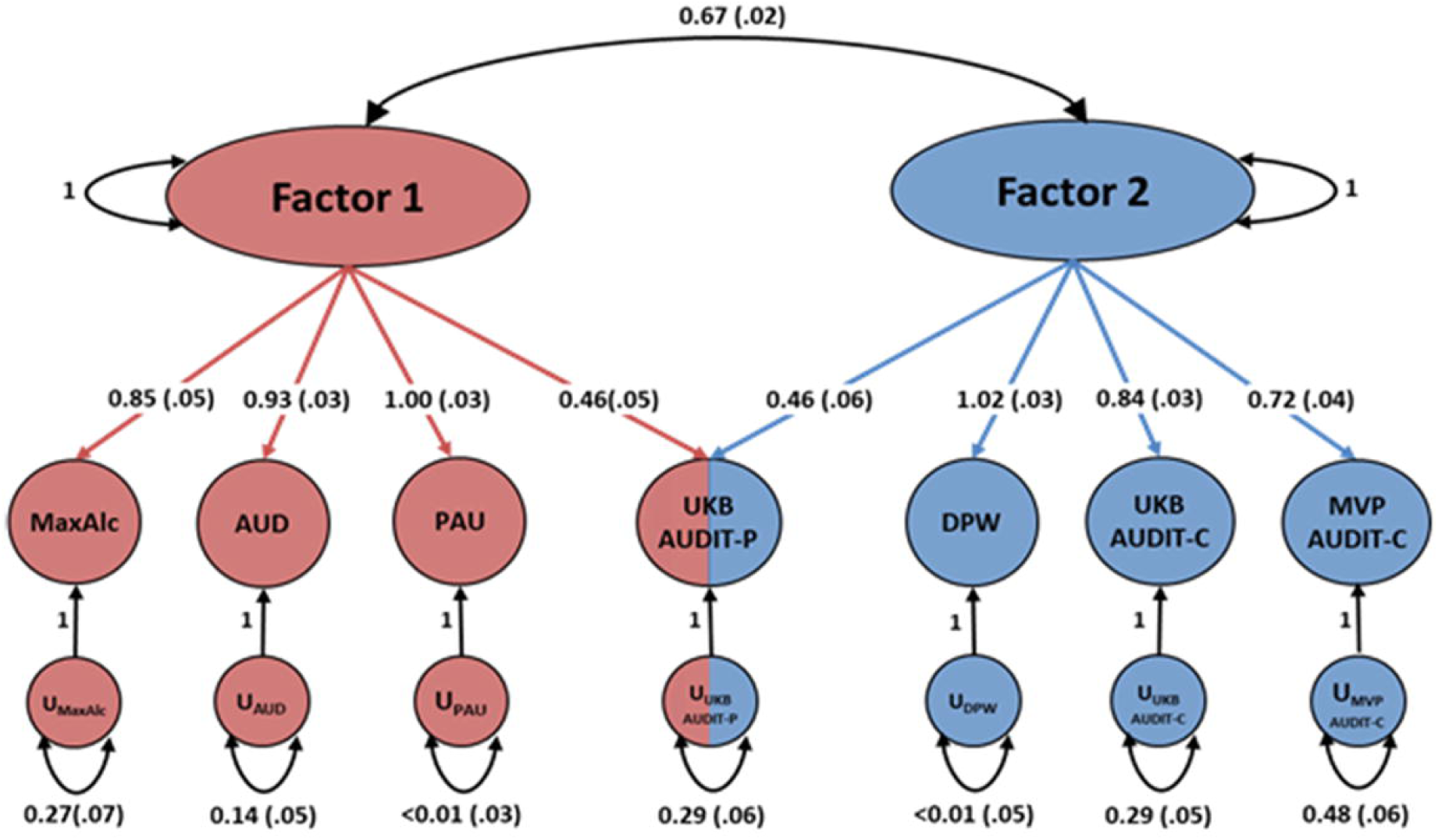
Genomic-SEM of MVP MaxAlc and other alcohol use traits. *Note:* Abbreviations (left to right): Maximum habitual alcohol intake (MaxAlc); Alcohol use disorder (AUD); Problematic alcohol use (PAU); Alcohol Use Disorders Identification Test-Problems (AUDIT-P); Drinks per week (DPW); AUDIT-Consumption (AUDIT-C). We interpret Factor 1 to capture “alcohol use problems” and Factor 2 to capture “non-dependent alcohol consumption.”

### Mendelian Randomization (MR)Analyses

Of the three liver enzyme levels examined (ALP, ALT, and GGT), only GGT was significantly genetically correlated with MaxAlc (r_g_=0.18; s.e.=0.03; *p*=1.17×10^−07^) (**Supplemental Table15**). We thus focused MR analyses on the relationship between MaxAlc and GGT. Following the removal of 13 genetic variant outliers identified by MR-PRESSO, IVW methods indicated a small, but statistically significant causal relationship between MaxAlc and GGT liver levels (beta=0.012; s.e.=0.002; *p*=2.66×10^−10^)(**Supplemental Table16**). MR-RAPs results indicated that the effect estimates were not biased by the inclusion of SNPs of suggestive significance (**Supplemental Table17)**. Even after accounting for all SNP outliers, unexplained heterogeneity remained, potentially due to demographic differences between the two datasets used for the MR analysis (**Supplemental Table16**).

### Multi-trait analysis of MaxAlc and PAU

The MaxAlc MTAG analysis provided an increase in EUR MaxAlc equivalent sample size from N=218,623 (GWAS mean L2=1.29) to an equivalent sample size of N=353,981 (MTAG mean L2=1.47), resulting in 31 independent GWS MaxAlc risk loci. The top association was with *ADH1B*\*rs1229984 (*p*=3.35×10^−176^) (**Supplemental Figure16; Table1**). Results for the MTAG analysis of PAU also showed increased locus discovery and are reported in **supplemental materials**.

## Discussion

These findings highlight the additional information that can be gleaned from GWAS of habitual alcohol consumption traits that augment prior conclusions from GWAS of other alcohol consumption measures. We identify GWS associations with both novel and well-replicated alcohol use loci (**Table2**) of etiological relevance for heavy alcohol use, demonstrate distinctive differences in the genetic architecture of AUD and MaxAlc from that of other alcohol consumption traits, and provide insight into the genetic etiology of MaxAlc and its relationship to other psychiatric and medical conditions.

**Table 2.**
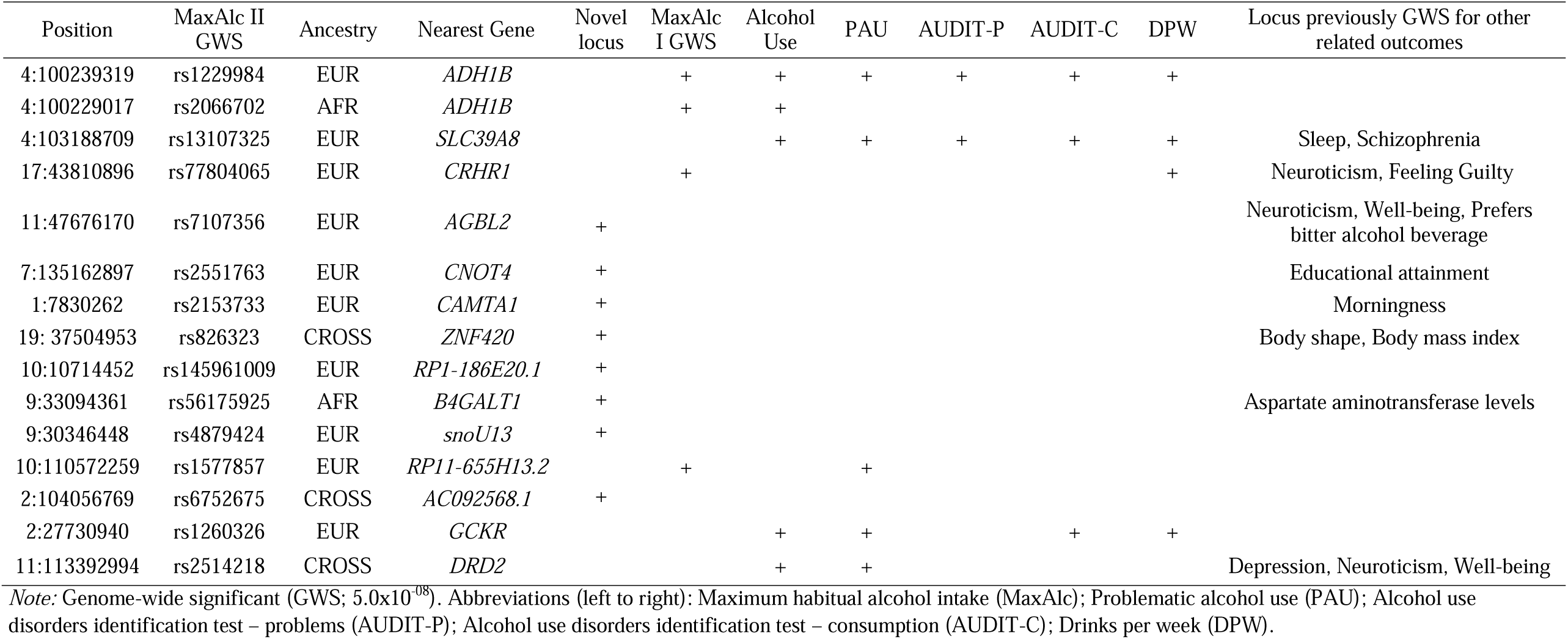
MVP MaxAlc loci summary table.

The strongest MaxAlc associations were with the two functional *ADH1B* loci— rs1229984 (*p*=3.12×10^−101^) in EUR and rs2066702 (*p*=6.30×10^−17^) in AFR. These *ADH1B* loci, among the few that survived from the candidate gene era(1,30-31), were also identified as risk loci for alcohol dependence in the respective ancestry groups in a GWS context(29). *ADH1B* has been associated with many alcohol traits studied using GWAS, including our previous MaxAlc study(8).

MaxAlc loci at *ADH1B, CRHR1* (rs77804065; *p*=6.04×10^−14^), and *RP11-655H13*.*2* (rs1577857; *p*=2.47×10^−08^) previously identified in EUR or AFR were confirmed in the present study, but the locus at *XPO7* (rs7821592; *p*=3.63×10^−08^), previously identified for MaxAlc in EUR(8), fell below GWS (*p*=1.21×10^−06^); although *XPO7* was GWS in the MaxAlc MTAG analysis (**Table1)**. Additional GWS associations included loci previously associated with alcohol consumption (3,5-6), PAU(4), and AUD(3) in *SLC39A8* (rs13107325; *p*=6.78×10^−15^), *GCKR* (rs1260326; *p*=4.53×10^−08^), and *DRD2* (rs2514218; *p*=4.59×10^−08^). A summary of GWS associations from the MaxAlc GWAS, including previous associations with other alcohol traits and etiologically relevant outcomes is presented **(Table2)**.

We also identified novel associations. For example, the MaxAlc EUR GWAS identified a GWS association (rs7107356; *p*=1.43×10^−10^) near *AGBL2* (AGBL Carboxypeptidase 2) on chromosome 11. *AGBL2* was previously identified as a risk locus for bitter alcoholic beverage taste preference(32). Bitter taste preference influences alcohol consumption(33), and genetic influences for bitter taste preference may influence alcoholic beverage intake(34).

A novel finding in the MaxAlc GWAS for AFR was the GWS association near *B4GALT1* (BETA-1,4, Galactrotransferase 1) on chromosome 9. *B4GALT1* is a member of the galactosyltransferase gene family and encodes an enzyme related to both glycoconjugate and lactose biosynthesis(35). Glycoconjugate metabolism occurring in the liver is often altered in the presence of chronic alcohol use, and glycoconjugate-related biomarkers are often considered markers of excessive alcohol use (differentiating alcohol-related vs non-alcohol-related tissue damage)(36). These findings show how MaxAlc GWAS can provide novel insight into the biology of alcohol use, including in non-EUR populations.

LDSC and gSEM results provide insight into differences and similarities in genetic architecture across a broad spectrum of alcohol consumption measures and AUD. Based on genetic correlations, MaxAlc is genetically more similar to AUD and PAU than to measures of alcohol consumption, and is distinct from less problematic drinking styles such as the UK Biobank measure of “having alcohol with meals.” MaxAlc, similar to AUD and PAU, demonstrated significant differences in genetic correlation with many psychiatric outcomes compared to the largely negative, or near zero, genetic correlations between psychopathology and other alcohol consumption measures. The strongest genetic correlation between MaxAlc and psychiatric traits was with suicide attempt, which is consistent with the positive phenotypic association between heavy alcohol consumption and suicide behaviors(37).

Our gSEM showed that a two-factor solution best fit the data, including one factor that we suggest captures “alcohol use problems” and a second factor that captures “non-dependent alcohol consumption.” MaxAlc loaded most strongly on the “alcohol use problems” factor along with AUD, PAU, and AUDIT-P (which loaded equally on both factors). Thus, MaxAlc, a measure of habitual alcohol consumption, aligns with traits that capture alcohol use problems rather than episode-limited measures of quantity or frequency of alcohol use. The genetic relationship between alcohol consumption and AUD thus appears dependent upon the steadiness and heaviness of alcohol consumption, which in turn appears to reflect dependent use.

The genetic study of MaxAlc is also positioned to study the impact of habitual alcohol use on alcohol-related disease, such as liver function. Understanding the causal role of problematic alcohol use on alcohol-related medical conditions is important. Previous studies have primarily been limited to examining the relationship between AUD diagnosis and disease(38) although we know that harmful levels of drinking can negatively impact health outcomes even if not meeting AUD diagnostic criteria(39). The MR findings from the current study supported significant genetic overlap and a small causal role of habitual drinking on GGT levels. Clinically, GGT elevations can be pronounced in individuals with a history of heavy drinking, individuals with alcohol-related cirrhosis, and can be valuable for detecting alcohol-induced liver disease(40). Genetically-informed studies of MaxAlc can help understand the biological and medical consequences of habitual drinking.

The current study has limitations. The MVP is a predominately male U.S. military Veteran sample and the results reflect this demographic. Additionally, retrospective reports such as MaxAlc may be subject to longitudinal changes and reporting bias(41). MaxAlc also has sample size restrictions for non-European subjects as is currently true for many genetic studies of complex traits.

The present study is valuable in understanding etiologically-relevant mechanisms involved as normative alcohol consumption approaches habitual problematic use, and ultimately, AUD(42). These efforts provide promise for the continued advancement of our understanding of the etiology across the spectrum of alcohol consumption levels and AUD, and how these levels of alcohol use relate to other psychiatric and health conditions.

## Data Availability

All data produced in the present study are available upon reasonable request to the authors

