## Supplemental Information for "Genome-wide investigation of maximum habitual alcohol intake (MaxAlc) in 247,755 European and African Ancestry U.S. Veterans informs the relationship between habitual alcohol consumption and alcohol use disorder"

*Genotyping, Imputation, and Quality Control*

Genotyping and imputation for MVP have been described previously(1-6). Briefly, genotyping was performed using a custom Affymetrix Axiom Array covering ~720,000 SNPs. Imputation for SNPs was then performed using MiniMac4(7) and an African Genome Resources (AGR) reference panel curated by the Sanger Institute. Indels and complex variants were imputed separately using the 1000 Genomes Project phase 3 (1kGP Phase 3) reference panel(8). Imputed variants with info score <0.30, genotype hard call missingness ≥0.20, and minor allele frequency <0.001 were removed. Eigenstrat(9) was used to conduct a principal components (PC) analysis to determine genetic ancestry using the 1kGP Phase 3 reference panel(8). MVP participants of European (EUR) and African (AFR) ancestry were identified by the first 10 PCs for inclusion in the current study.

*Gene, Gene-set, and Tissue-Specific Gene Expression Analysis*

FUMA (Functional Mapping and Annotation)(10) was used to conduct gene-based, gene-set, and tissue-specific gene expression analyses using MAGMA (Multi-marker Analysis of GenoMic Annotation)(11). SNPs were mapped to 18,723 genes for EURs and 19,024 for AFRs. Ancestry-specific Bonferroni corrections were used to determine GWS (EUR: *p*=0.05/18,723=2.67x10^-06^; AFR: *p*=0.05/19,024 = 2.63x10^-06^) for gene-based tests. MAGMA was also used to conduct analyses of gene sets classified based on gene function and biological processes (MsigDB). Tissue-specific gene expression was also analyzed using tissue transcriptomic profile data from GTEx v8(12).

In EUR, 27 genes reached Bonferroni-corrected GWS (*p*=2.67x10^-06^) in the gene-based test (**Supplemental Figure4; Supplemental Table2**). The top gene-based association was with *KANSL1* (*p*=8.67x10^-14^) on chromosome 17. Thirteen additional genes, including *CRHR1*, map in close proximity to *KANSL1* in a well-known inversion region on chromosome 17 and were also GWS. No gene sets were GWS in the EUR analysis. Multiple brain regions were enriched in the EUR tissue-expression analysis including cerebellum (*p*=8.25x10^-06^), cerebellar hemisphere (*p*=8.46x10^-06^), frontal cortex (*p*=5.05x10^-05^), brain cortex (*p*=2.19x10^-04^), anterior cingulate cortex (*p*=2.19x10^-04^), hypothalamus (*p*=2.36x10^-04^), and the nucleus accumbens basal ganglia (*p*=5.46x10^-04^). (**Supplemental Figure5; Supplemental Table4**).

In AFR, *ADH1B* (*p*=3.28x10^-08^) and *METAP1* (*p*=7.11x10^-07^) on chromosome 4 were GWS in the gene-based test (**Supplemental Figure 6)**. The AFR gene-set analysis resulted in two GWS gene sets: GO_bp:go_retinoic_acid_metabolic_process (*p*=2.62x10^-08^*;p_bonferr_*=4.05x10^-04^) and GO_mf:go_alcohol_dehydrogenase_activity_zinc_dependent (*p*=1.95x10^-07^*;p_bonferr_*=3.01x10^-03^)(**Supplemental Table6)**. There were no significant tissue expression findings in the AFR MaxAlc analysis.

*Functional characterization of identified genetic risk loci*

Genetic variants were characterized using two gene-mapping approaches: (1) by using expression quantitative trait locus (eQTL) data from GTEx v8(12) and BRAINEAC(13); and (2) using 3D chromatin interactions (Hi-C)(14). Both approaches were implemented in the FUMA platform (Functional Mapping and Annotation)(10).

Gene-mapping using eQTL data was performed using GTEx v8 gene expression data including gene expression data for: amygdala, anterior cingulate cortex BA24, cerebellar hemisphere, cerebellum, cortex, frontal cortex BA9, hippocampus, hypothalamus, nucleus accumbens basal ganglia, putamen basal ganglia, spinal cord cervical c-1, and substantia nigra(12). BRAINEAC(13) tissues included: cerebellar cortex, frontal cortex, hippocampus, inferior olivary nucleus, occipital cortex, putamen, substantia nigra, temporal cortex, thalamus, and intralobular white matter. Hi-C chromatin interaction data included PsychENCODE(15) EP links and PsychENCODE promoter anchored loops, and FUMA-based datasets for Hi-C in adult cortex, dorsolateral prefrontal cortex, and hippocampus. Circos plots for chromosomes containing genome-wide significant MaxAlc loci are presented in **Supplementary Figure18-19**.

*Multi-trait analysis of MaxAlc and Problematic alcohol use (PAU)*

A multi-trait analysis of GWAS (MTAG)(16) was performed using summary statistics from the EUR MaxAlc GWAS and the previously published GWAS of PAU(17). MaxAlc and PAU were strongly genetically correlated (rg=0.79). MTAG leverages the high degree of genetic correlation between related traits to generate trait-specific effect estimates for each genetic variant, and thus, can enhance statistical power for trait-specific genetic discovery through the inclusion of multiple GWAS summary statistics while also accounting for any sample overlap(16).

The MTAG analysis was restricted to SNPs in common to both the EUR MaxAlc GWAS (N=218,623) and the PAU GWAS (N_effective_=300,789), with a minor allele frequency > 0.01, and occurring in at least 50% of the effective sample size of the GWAS summary statistics. Because information from both sets of GWAS summary statistics are informative for the other included trait, effectively serving to boost the power of each respective study through the inclusion of the other, MTAG generates both MaxAlc-specific and PAU-specific effect estimates for all included SNPs. Thus, MTAG results are presented for both the MaxAlc trait and the PAU trait.

The top association in the MaxAlc MTAG analysis was with *ADH1B* rs1229984 (*p*=3.35x10^-176^). Many additional variants of interest were also identified, including the *XPO7* gene (rs1484162; *p*=1.49x10^-08^) that was genome-wide significant in the initial MaxAlc GWAS(2); however, dropped below GWS in the EUR MaxAlc GWAS in the current study (**Supplemental Figure16; Table 1; Supplemental Table18 [includes MTAG effect estimates]**).

The MTAG analysis for PAU resulted in a jump in sample size from a maximum effective sample size of 300,789 (GWAS mean ꭓ^2^=1.35) to an equivalent sample size of N=422,491 (MTAG mean ꭓ^2^=1.49). The PAU-specific MTAG resulted in 42 independent GWS PAU risk loci. The top PAU association was also with *ADH1B* rs1229984 (*p*=2.38x10^-182^) (**Supplemental Figure17; Supplemental Table19**).
