## Supplemental Figures for "Genome-wide investigation of maximum habitual alcohol intake (MaxAlc) in 247,755 European and African Ancestry U.S. Veterans informs the relationship between habitual alcohol consumption and alcohol use disorder"

**Supplemental Figure 1. Screenshot of the MVP MaxAlc survey item captured from Million Veteran Program Lifestyle Survey (Veterans Health Administration R&D; 2011).**

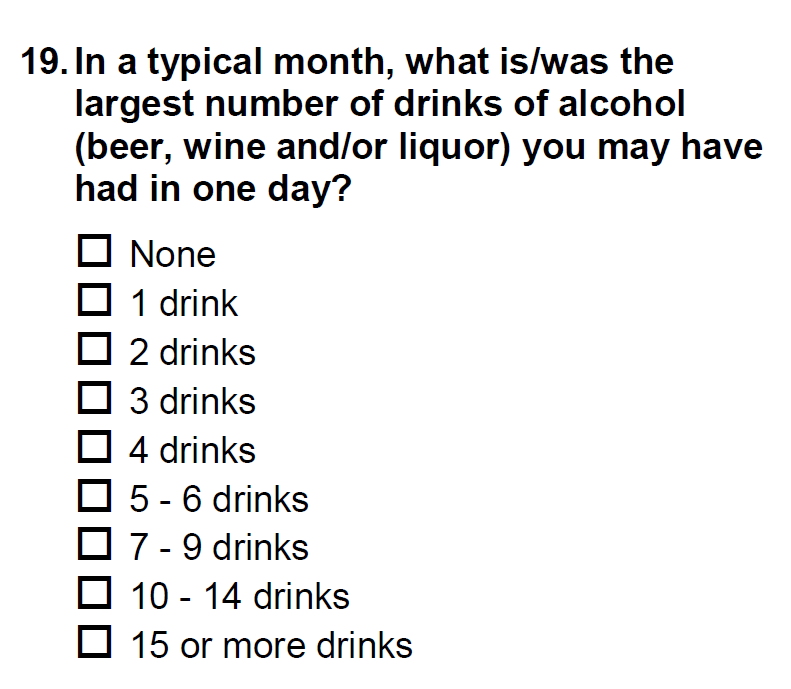

**Supplemental Figure 2. Distribution of MVP participant’s MaxAlc survey item responses: (a) all included participants; (b) AFR participants only; (c) EUR participants only.**

**(a)**

|  |  |
| --- | --- |
| None | 14177 |
| 1 drink | 31736 |
| 2 drinks | 47867 |
| 3 drinks | 36237 |
| 4 drinks | 28120 |
| 5-6 drinks | 35892 |
| 7-9 drinks | 17510 |
| 10-14 drinks | 14620 |
| 15 or more drinks | 21596 |

**(b)**

|  |  |
| --- | --- |
| None | 2436 |
| 1 drink | 3399 |
| 2 drinks | 5259 |
| 3 drinks | 4364 |
| 4 drinks | 3649 |
| 5-6 drinks | 4314 |
| 7-9 drinks | 1929 |
| 10-14 drinks | 1460 |
| 15 or more drinks | 2322 |

**(c)**

|  |  |
| --- | --- |
| None | 11741 |
| 1 drink | 28337 |
| 2 drinks | 42608 |
| 3 drinks | 31873 |
| 4 drinks | 24471 |
| 5-6 drinks | 31578 |
| 7-9 drinks | 15581 |
| 10-14 drinks | 13160 |
| 15 or more drinks | 19274 |

**Supplemental Figure 3. Manhattan plot of EUR ancestry MaxAlc GWAS: 10 independent genome-wide significant (GWS) loci.**

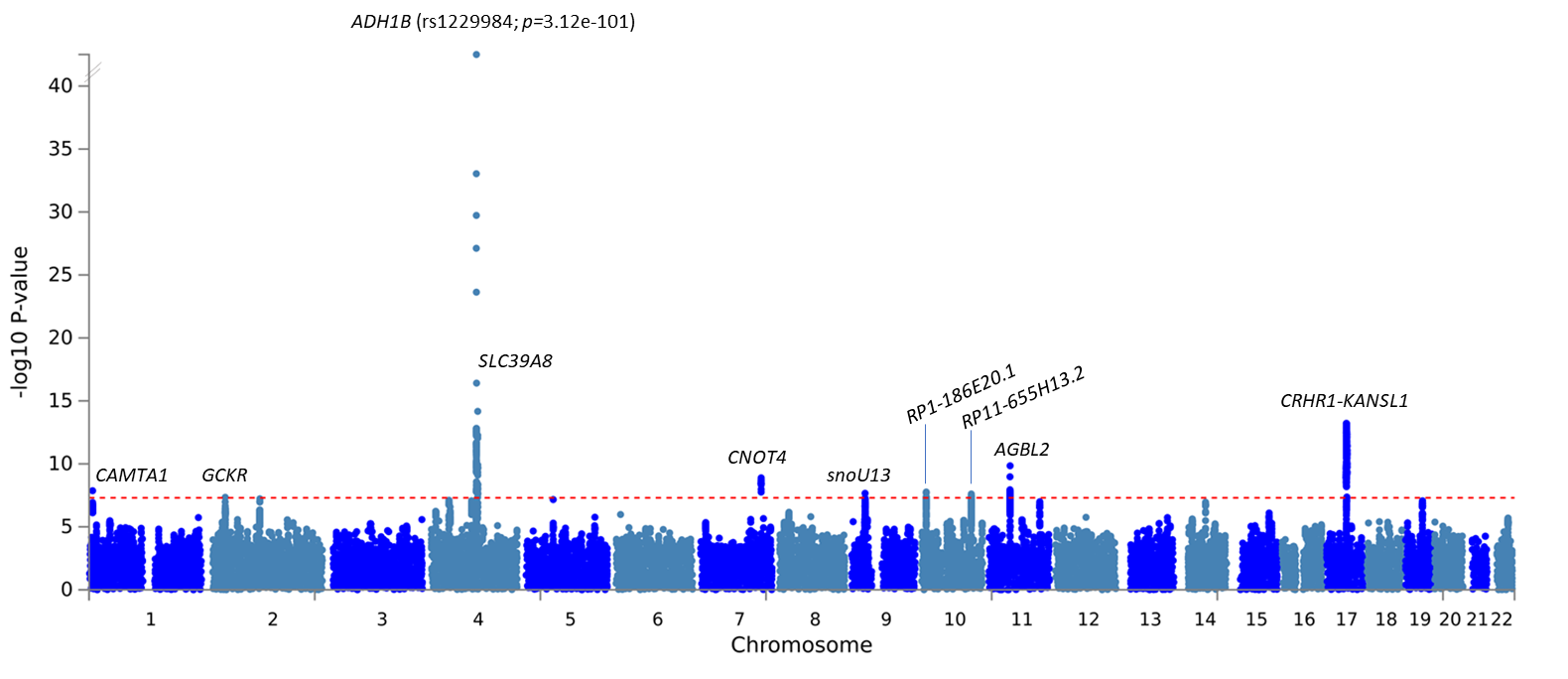

**Supplemental Figure 4. Manhattan plot of EUR ancestry MaxAlc gene-based results.**

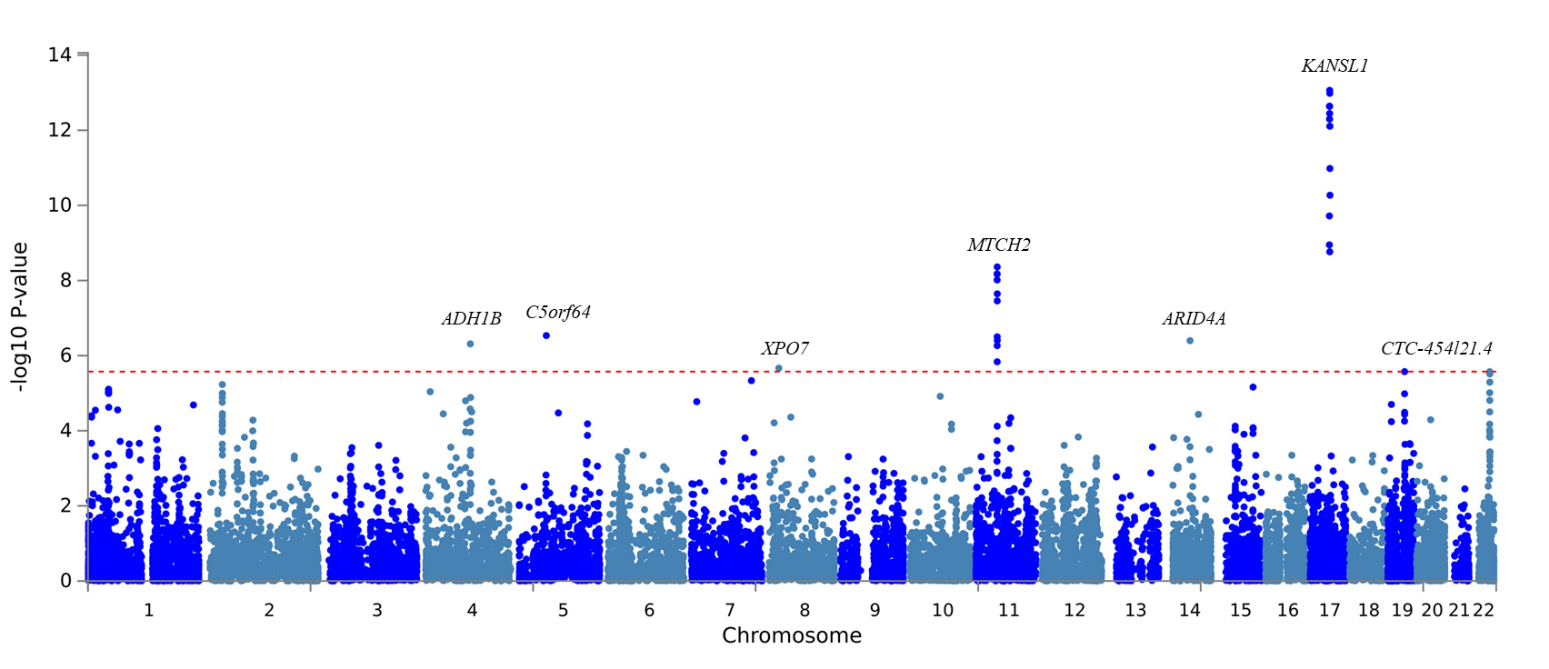

**Supplemental Figure 5. Tissue enrichment in EUR ancestry MaxAlc analysis.**

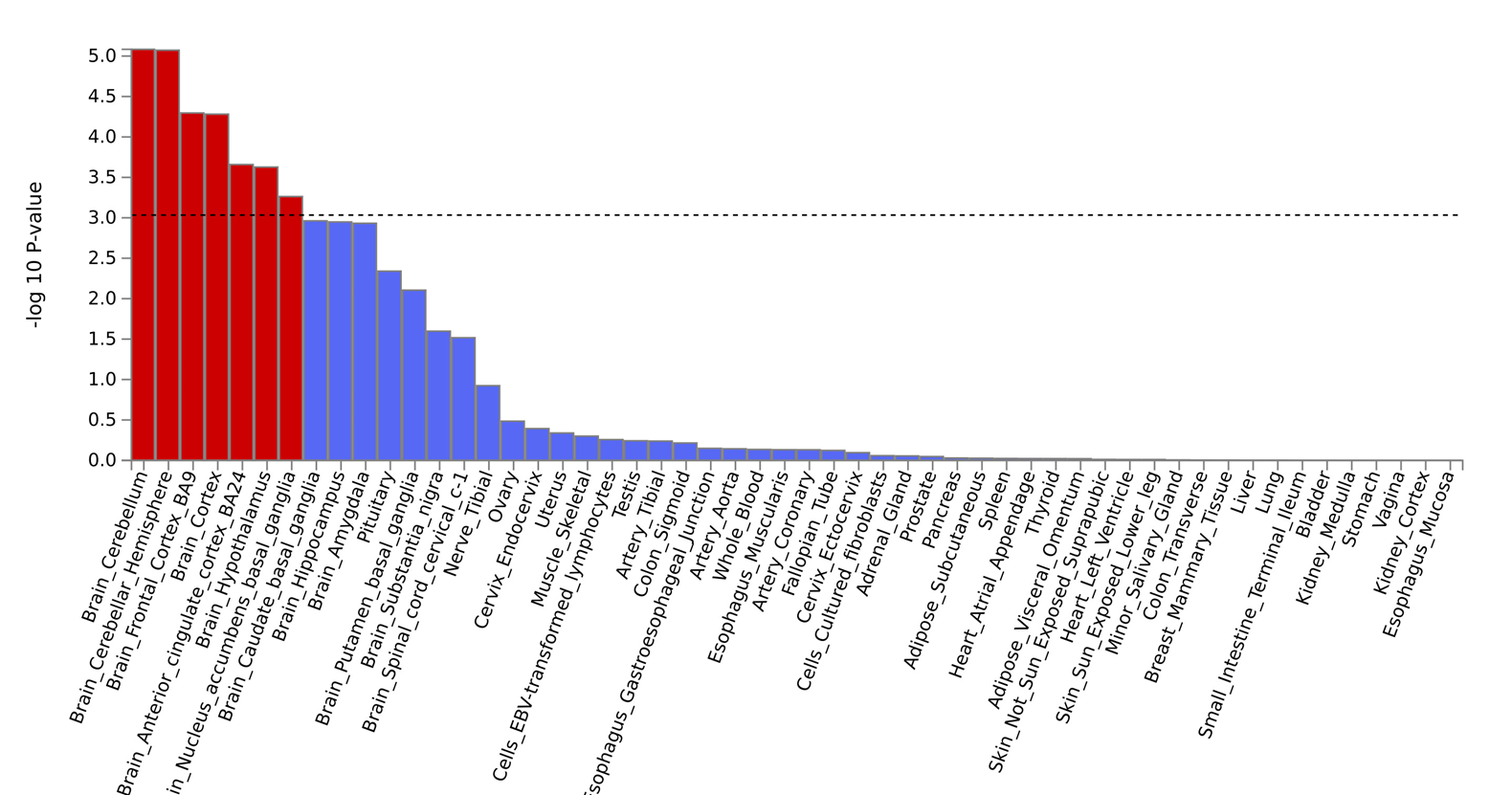

**Supplemental Figure 6. Manhattan plot of AFR ancestry MaxAlc GWAS: 2 independent genome-wide significant (GWS) loci.**

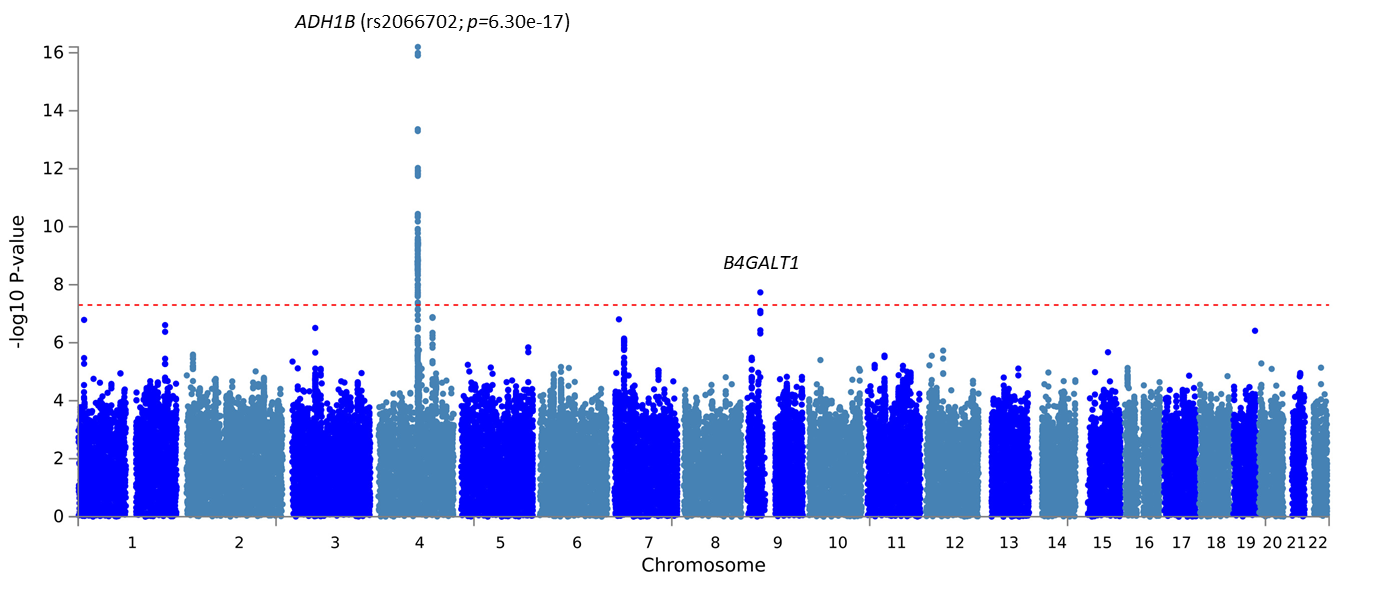

**Supplemental Figure 7. Manhattan plot of AFR ancestry MaxAlc gene-based results.**

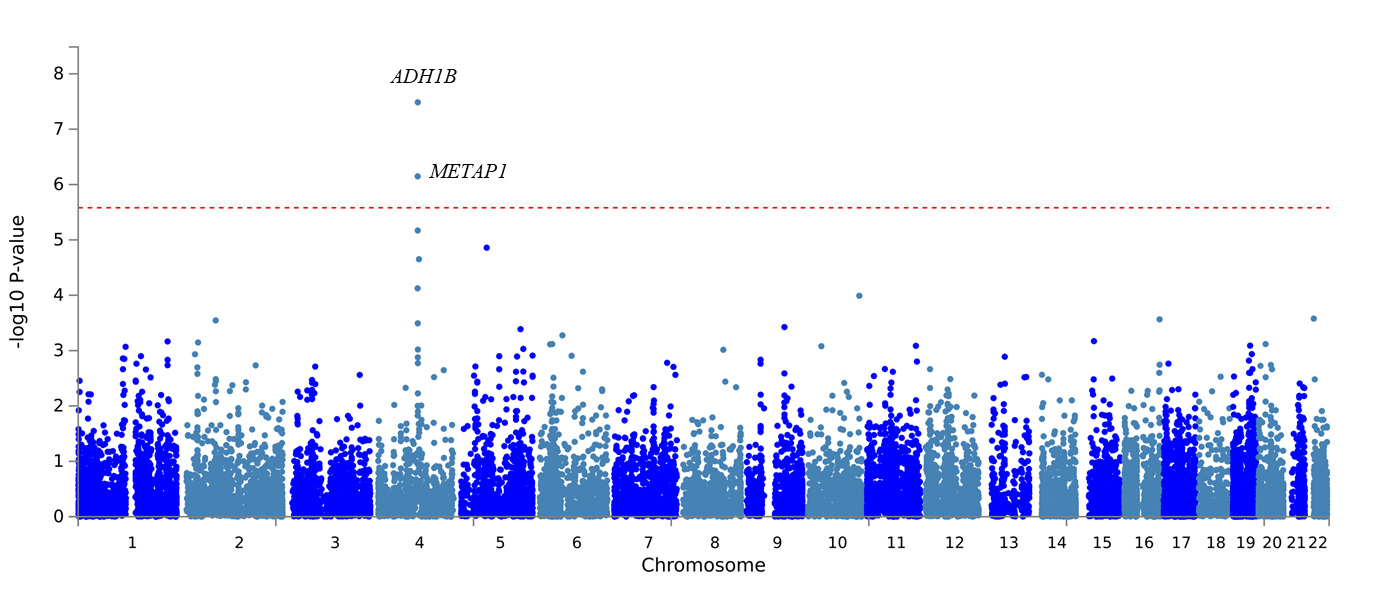

**Supplemental Figure 8. Tissue enrichment in AFR ancestry MaxAlc analysis.**

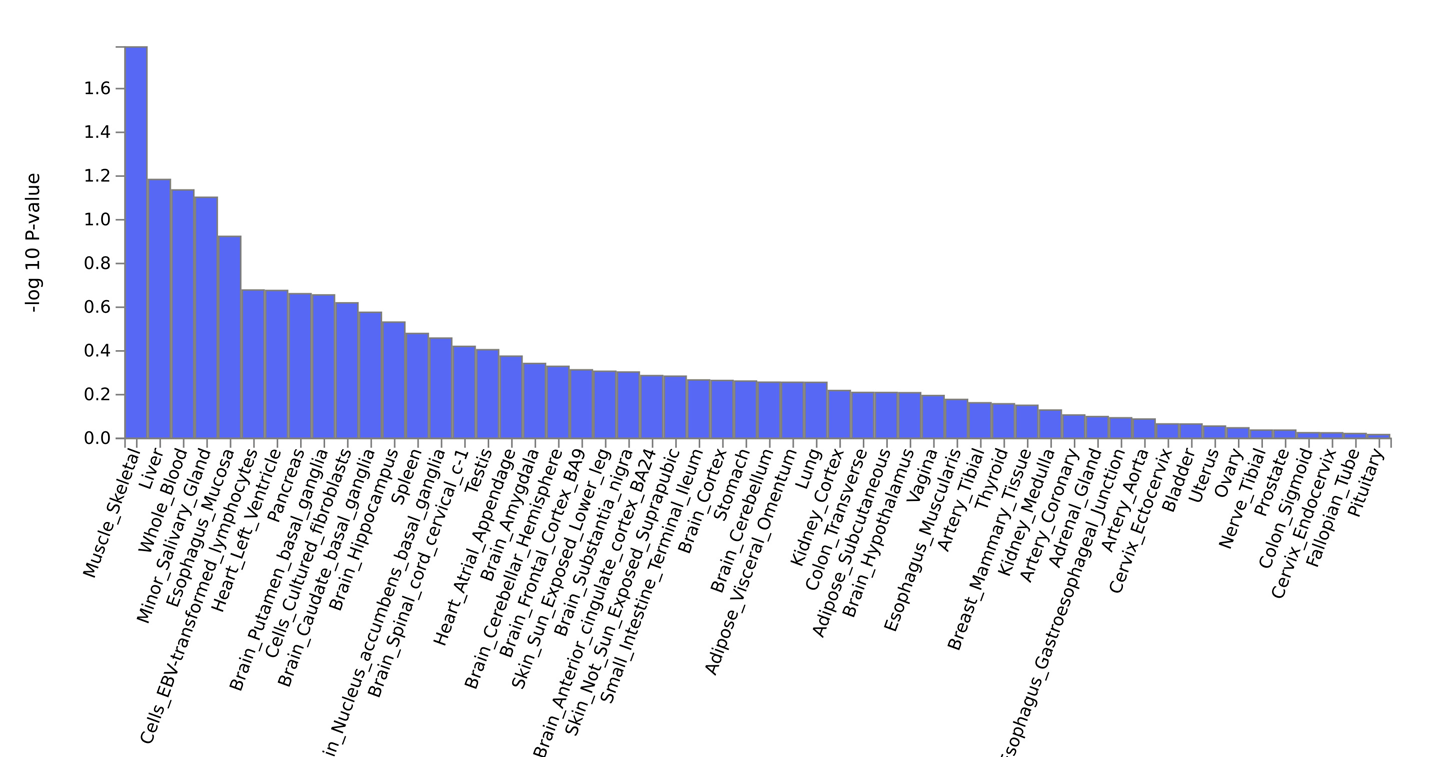

**Supplemental Figure 9. Manhattan plot of cross-ancestry MaxAlc gene-based results.**

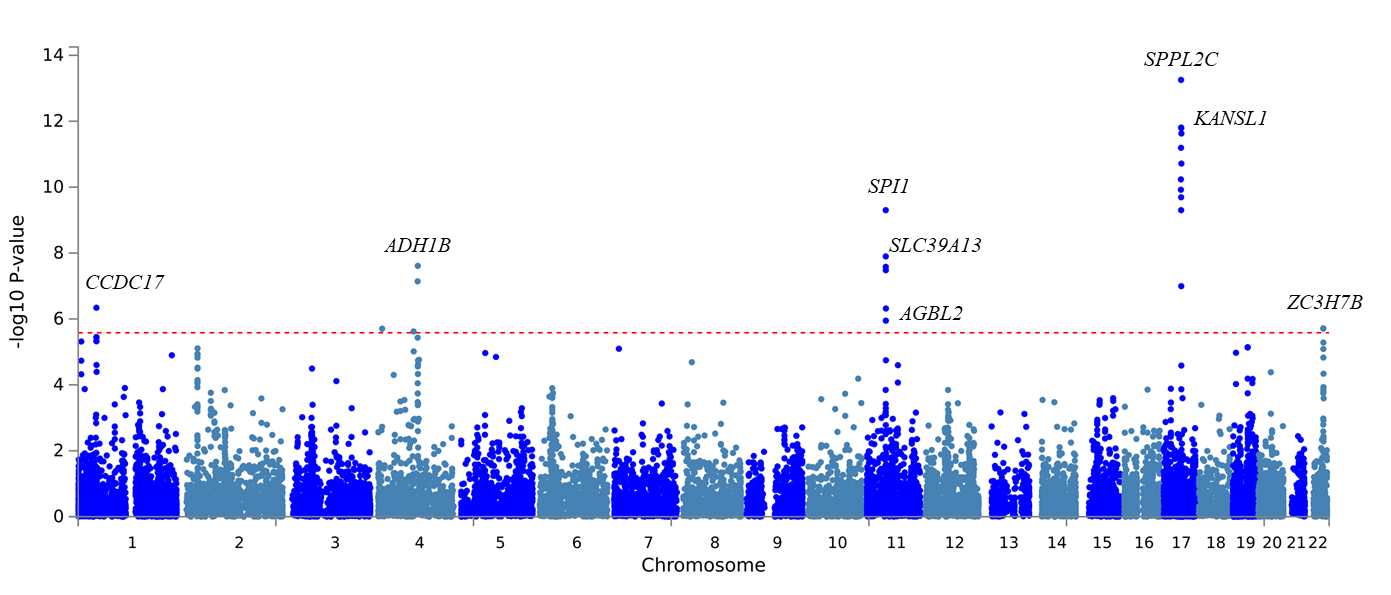

**Supplemental Figure 10. Tissue enrichment in cross-ancestry MaxAlc analysis.**

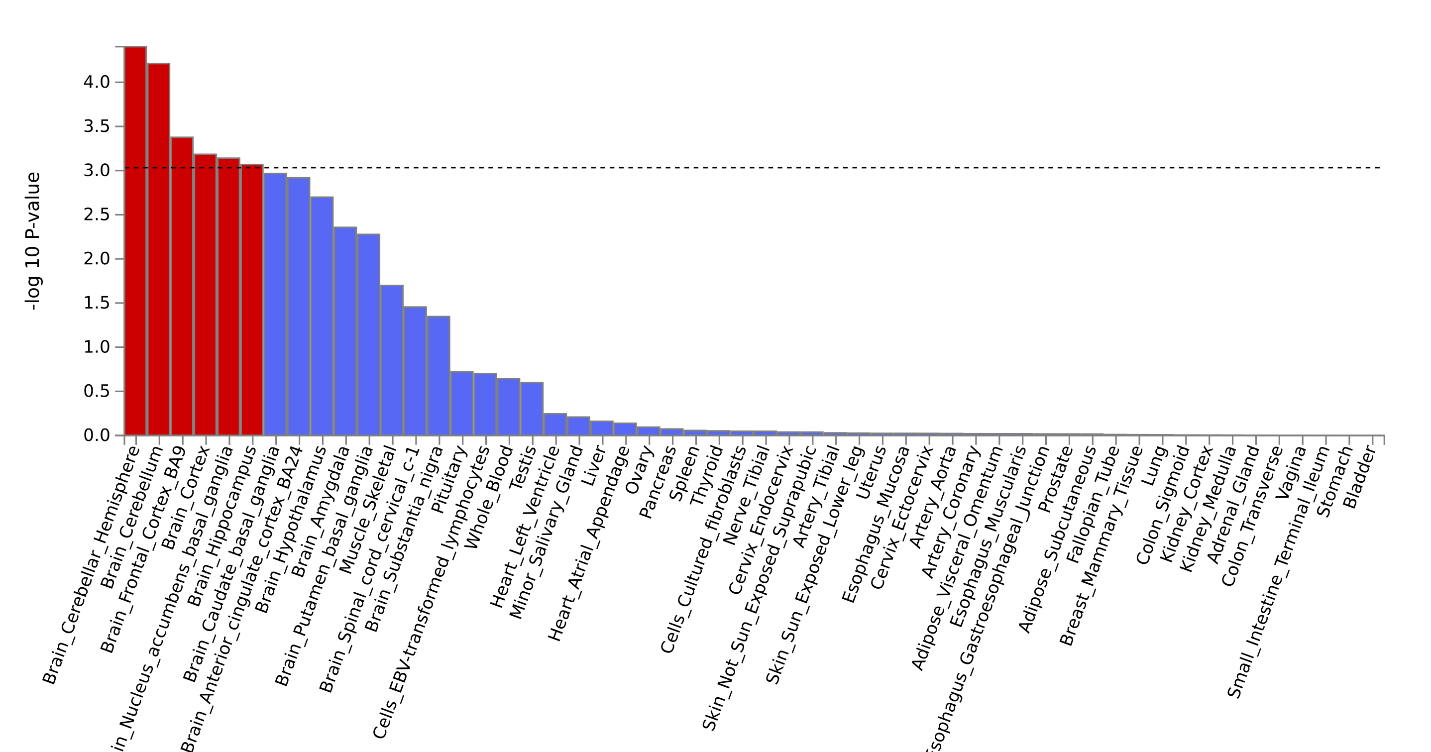

**Supplemental Figure 11. Mendelian Randomization – Scatter Plot**

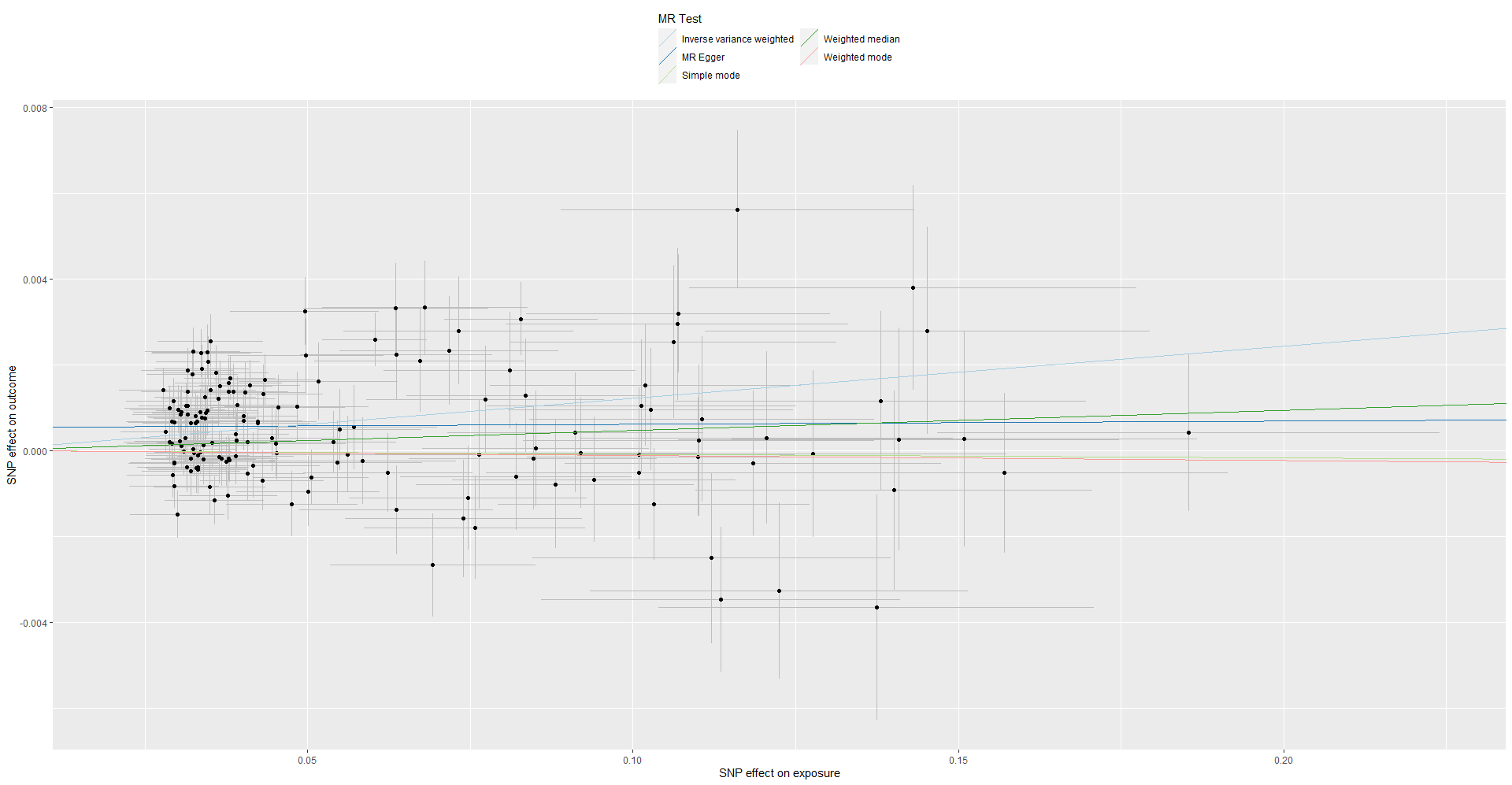

**Supplemental Figure 12. Mendelian Randomization – Forest Plot**

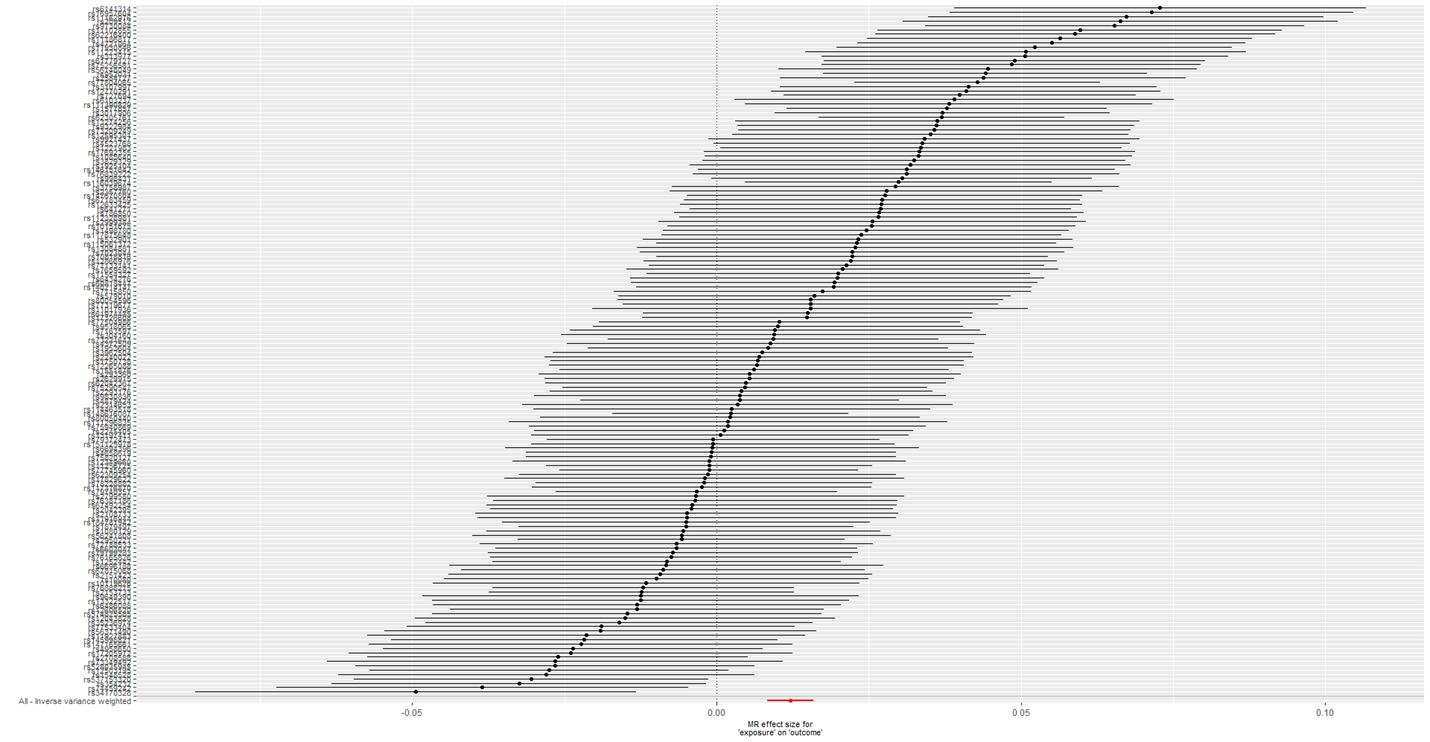

**Supplemental Figure 13. Mendelian Randomization – Leave-one-out Plot**

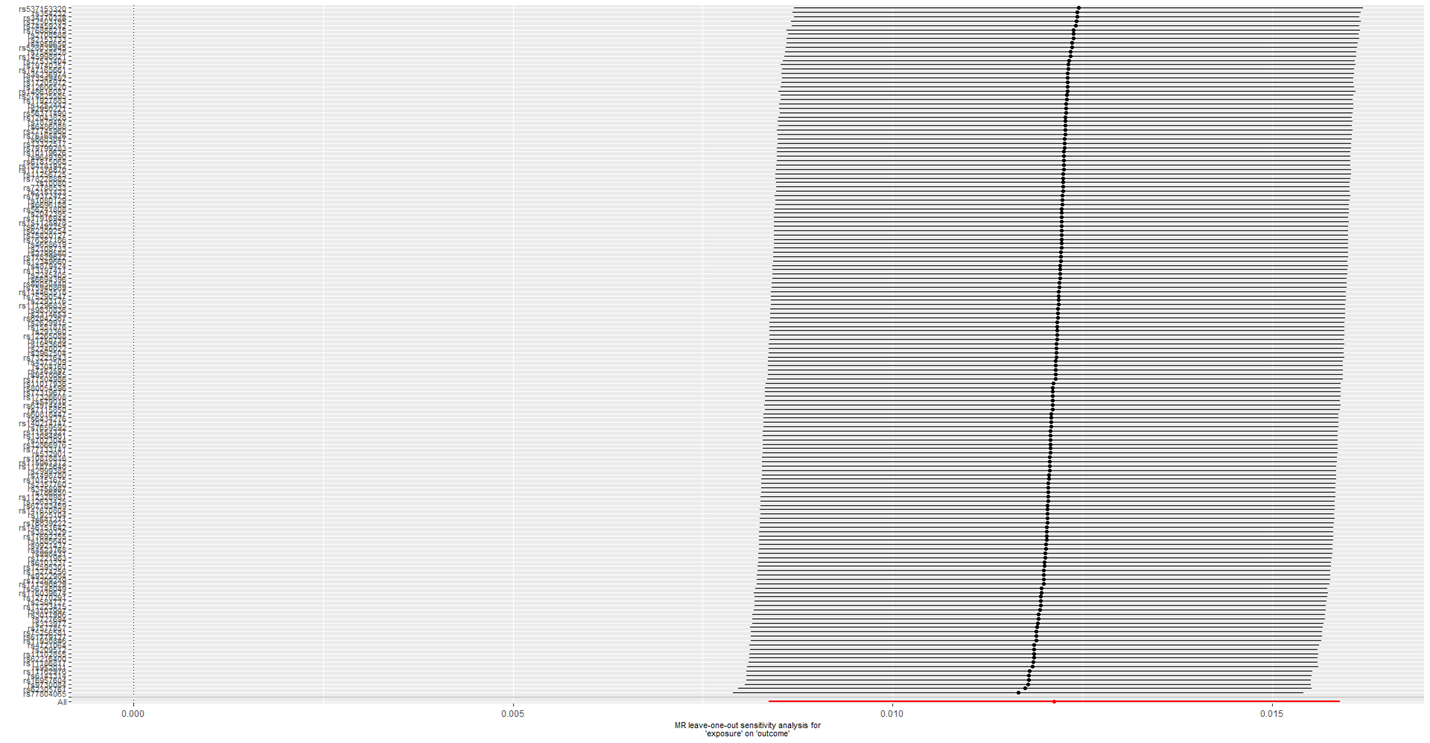

**Supplemental Figure 14. Mendelian Randomization – Funnel Plot**

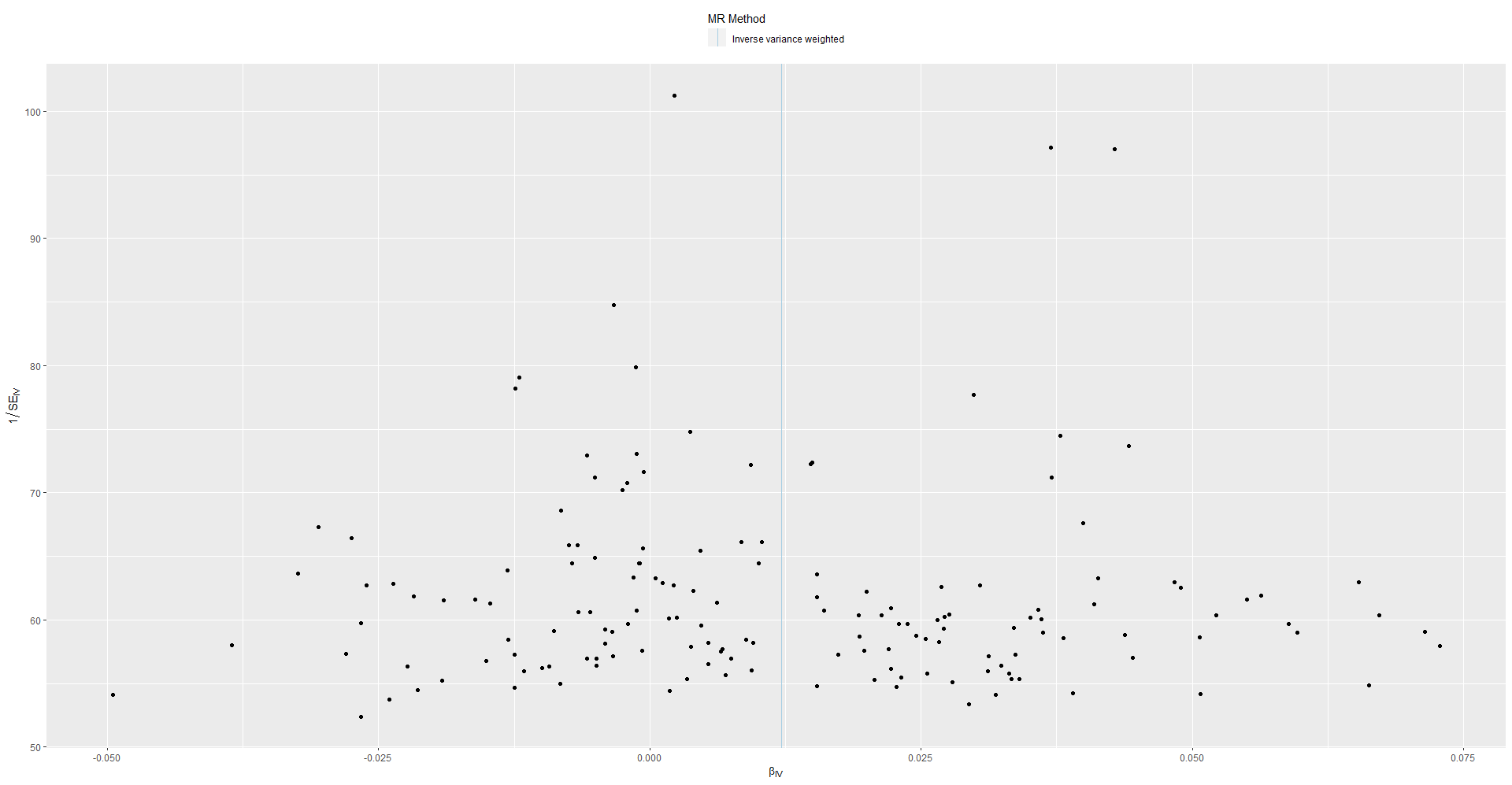

**Supplemental Figure 16: Manhattan plot of MaxAlc MTAG GWAS results (MaxAlc-PAU): 31 independent genome-wide significant (GWS) loci.**

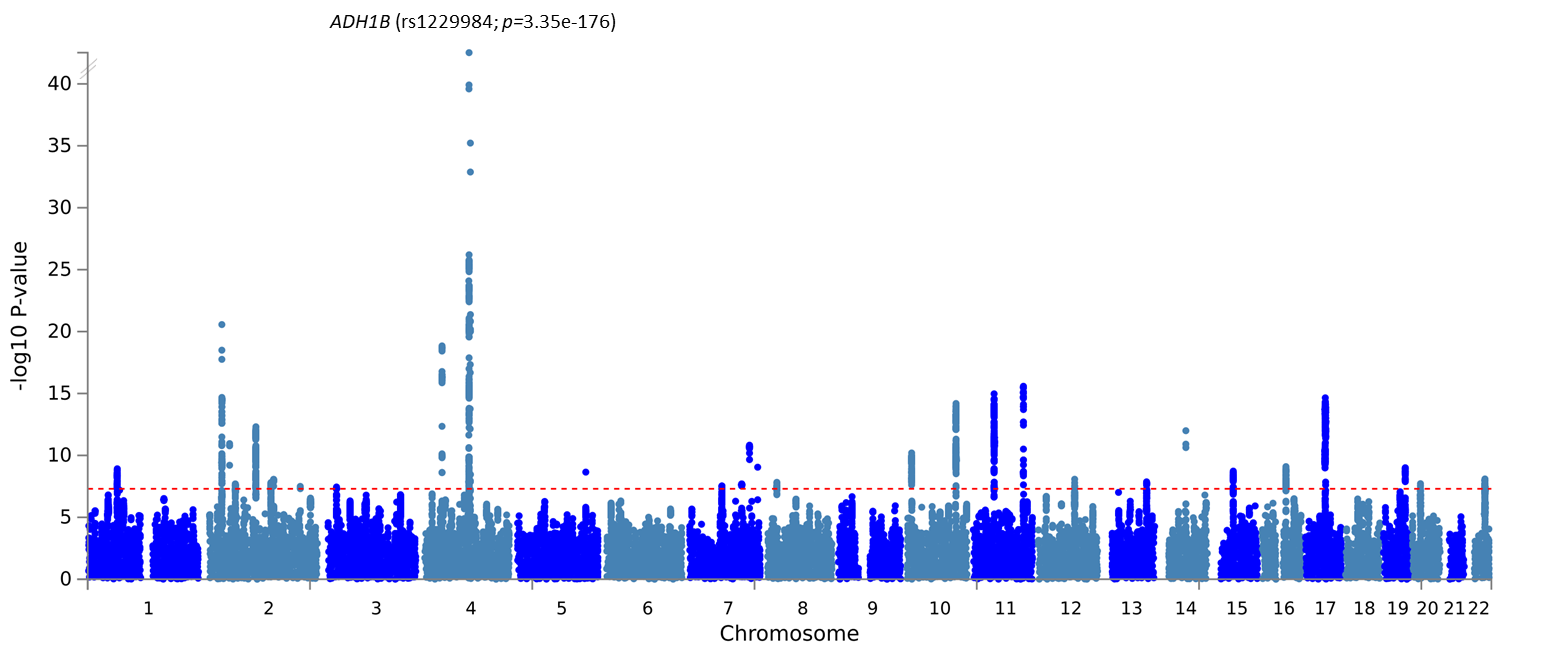

**Supplemental Figure 17: Manhattan plot of PAU MTAG GWAS results (PAU-MaxAlc): 42 independent genome-wide significant (GWS) loci.**

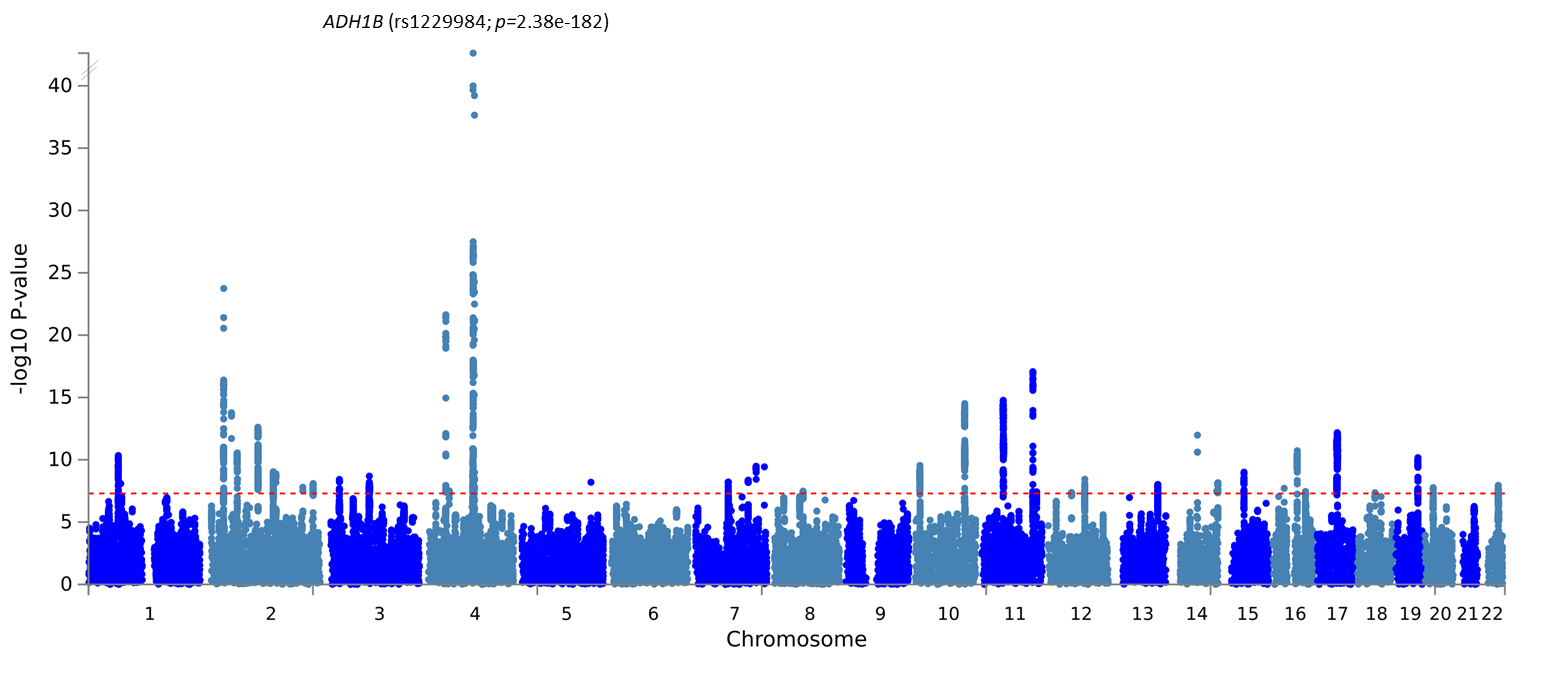

**Supplemental Figure 18: Circos plots for chromosomes containing genome-wide significant loci for the MaxAlc GWAS.**

**Note. Outer most layer** is a Manhattan plot of genome-wide association study (GWAS) single-nucleotide polymorphisms (SNPs) with *p*≤0.05. SNPs are plotted by chromosomal position along the *x*-axis with their corresponding -log^-10^ *p*-value on the *y*-axis. Linkage-disequilibrium (LD) between the identified lead SNP and surrounding SNPs is indicated from r^2^>0.8 (red), r^2^>0.6 (orange), r^2^>0.4 (green), r^2^>0.2 (blue). SNPs that are not in LD with the lead SNP (r^2^≤0.02) are gray. **Second layer (chromosome ring):** Chromosomal regions containing identified genomic risk loci are colored in blue. The names of genes implicated based upon variant associations with brain tissue expression quantitative trait loci (eQTLs) are colored green. The names of genes implicated based upon 3D chromatin interactions (Hi-C) are colored orange. Genes that are mapped based upon both eQTLs and Hi-C associations are colored red. **Third layer (chromosome ring):** Variants mapped to genes based upon associations with brain tissue eQTLs are linked in green. Variants mapped to genes based upon Hi-C data are linked in orange.

(a) **Chromosome 1**

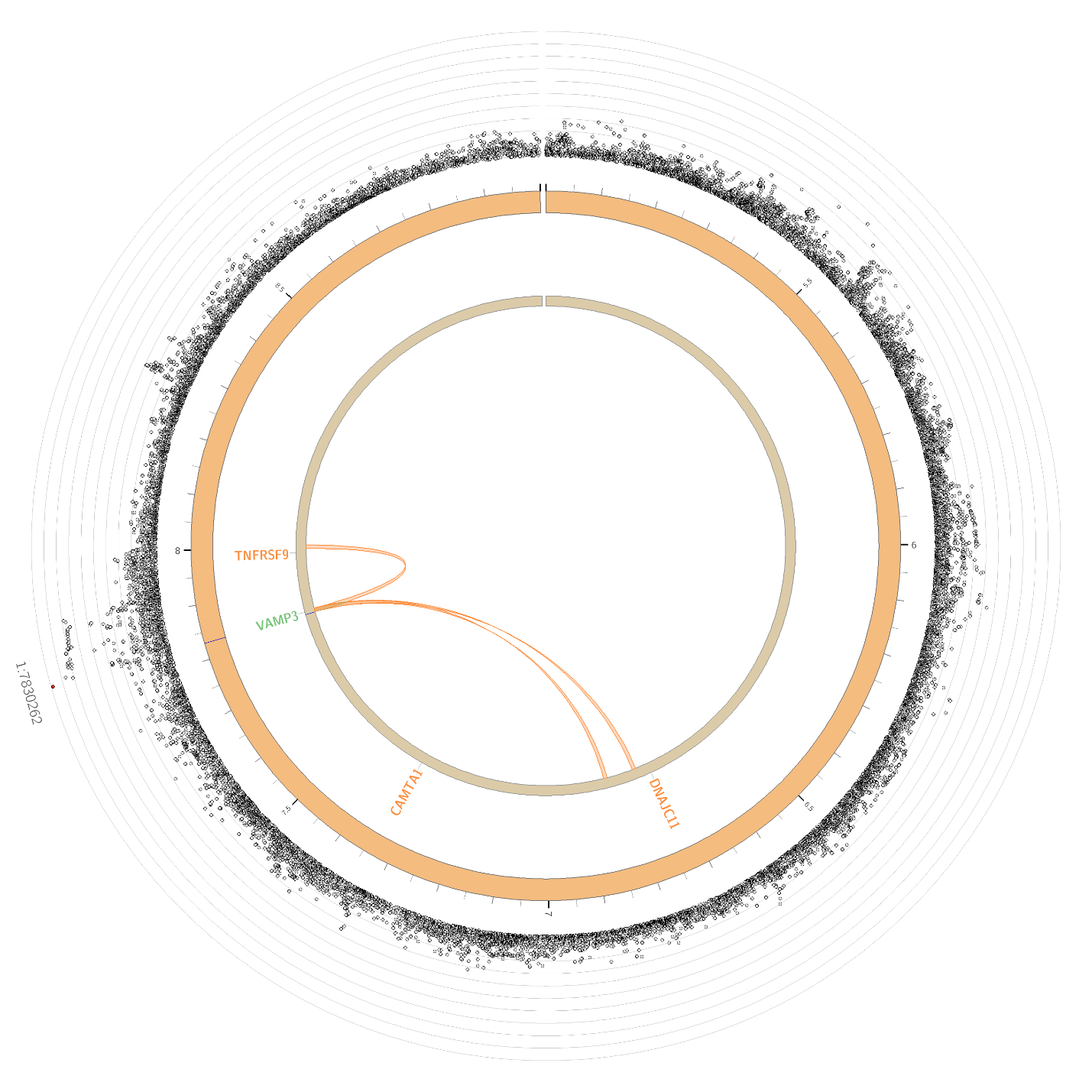

(b) **Chromosome 2**

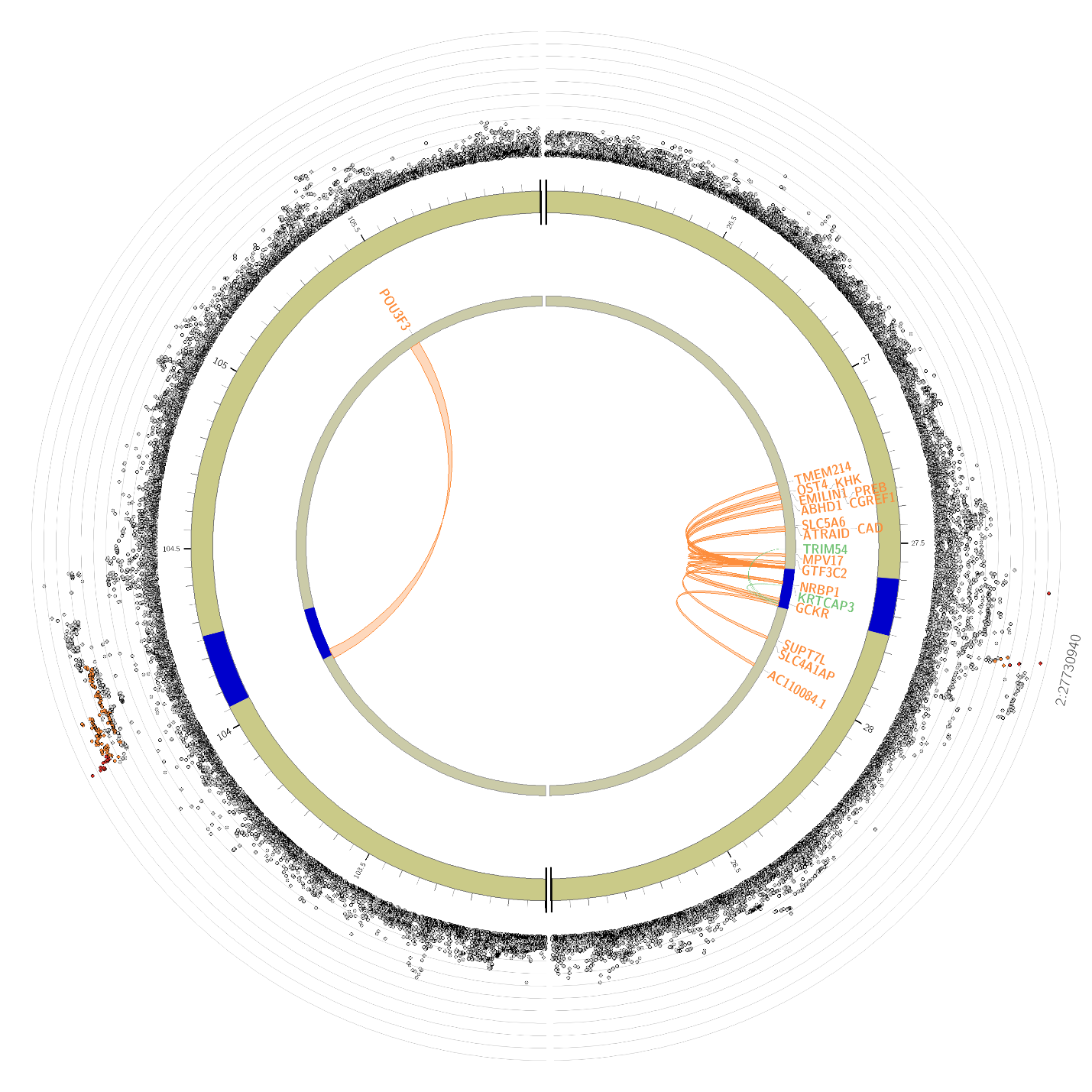

(c) **Chromosome 4**

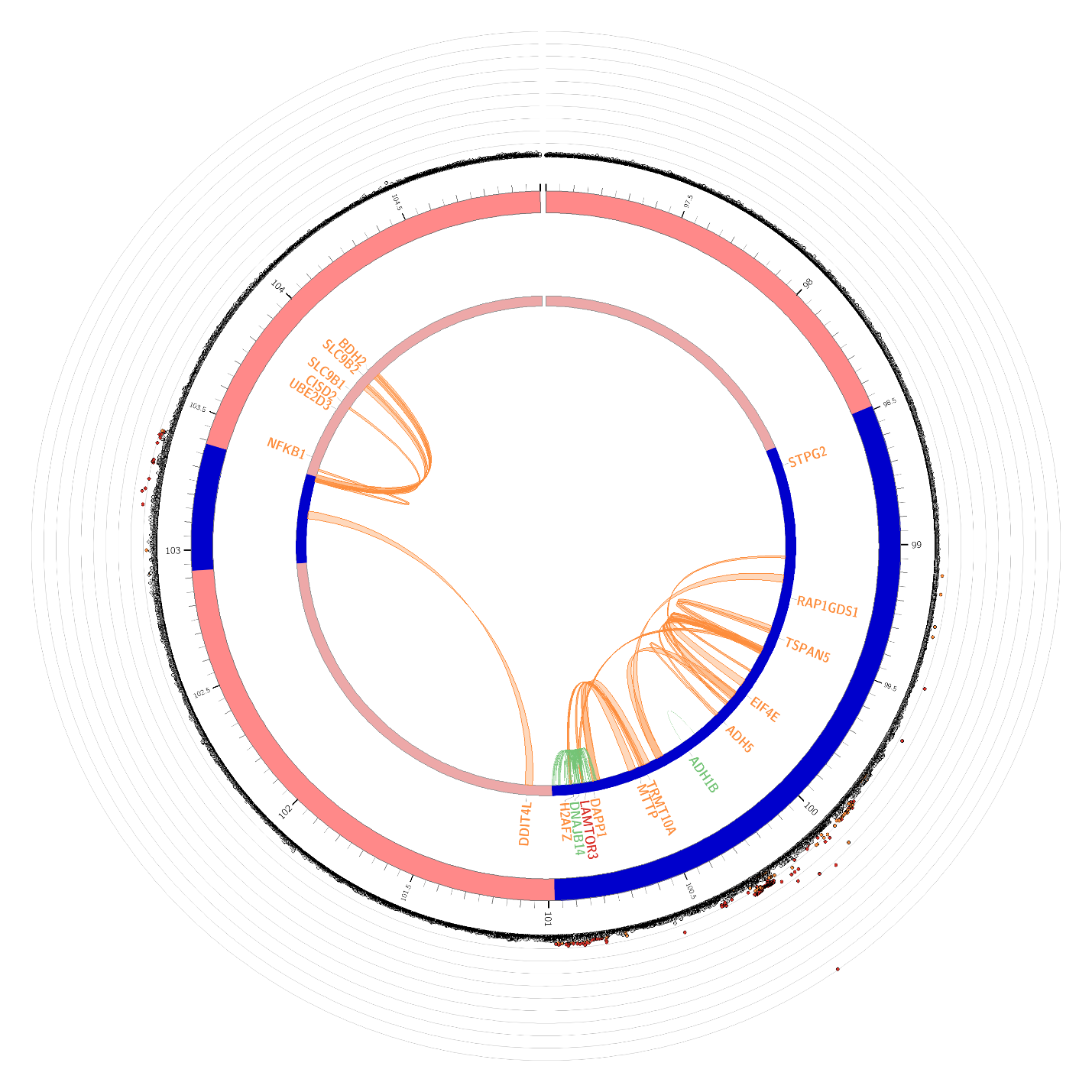

(d) **Chromosome 7**

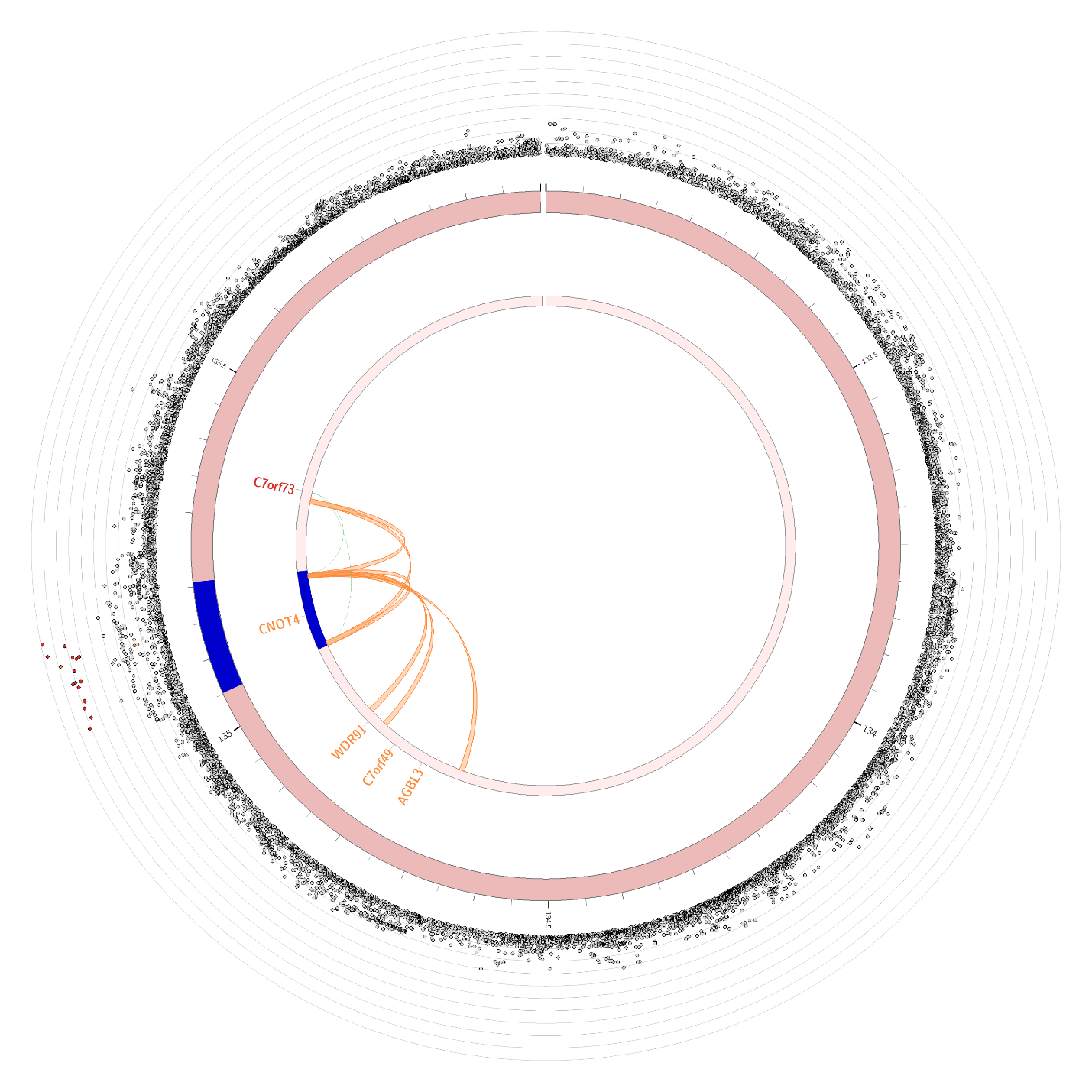

(e) **Chromosome 9**

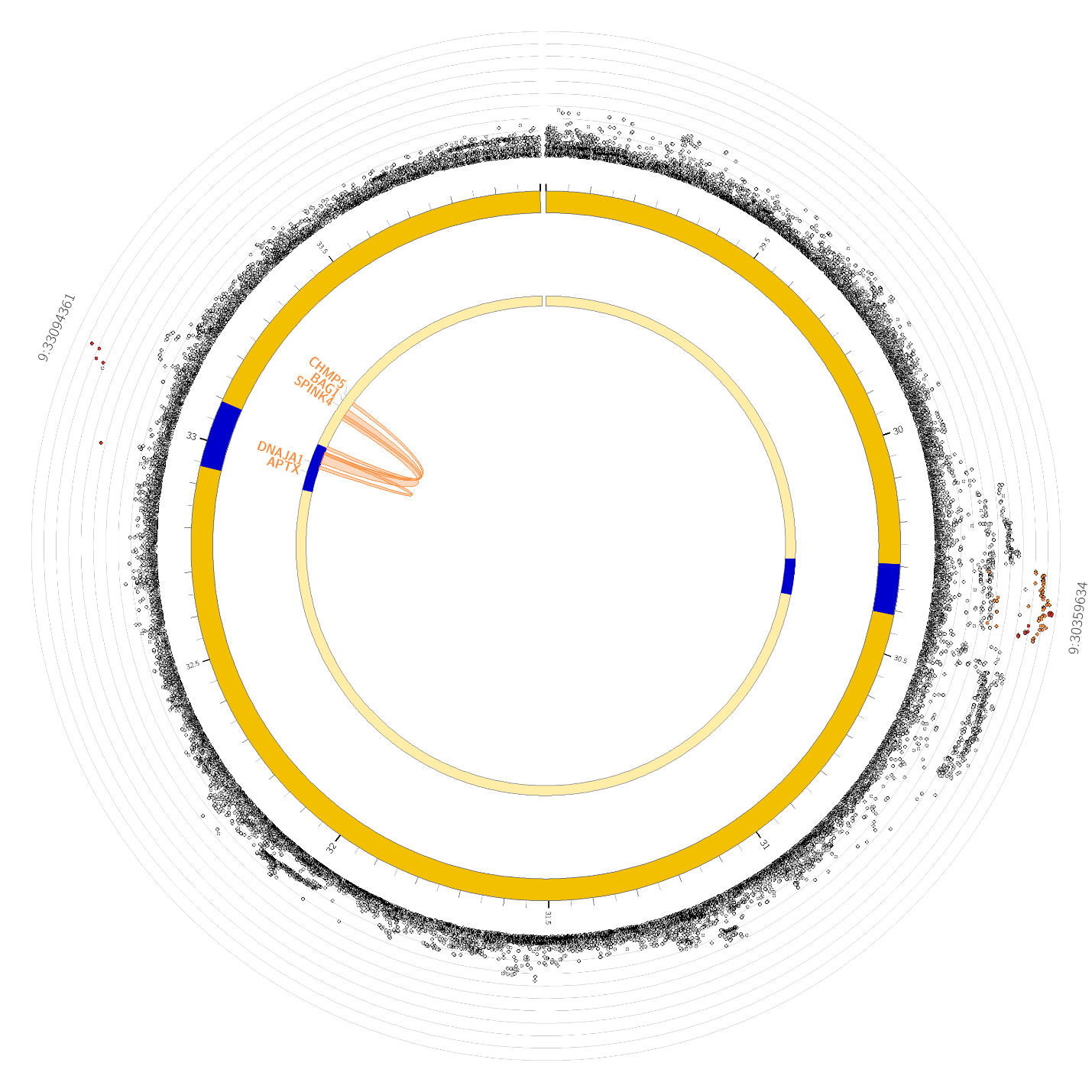

(f) **Chromosome 10**

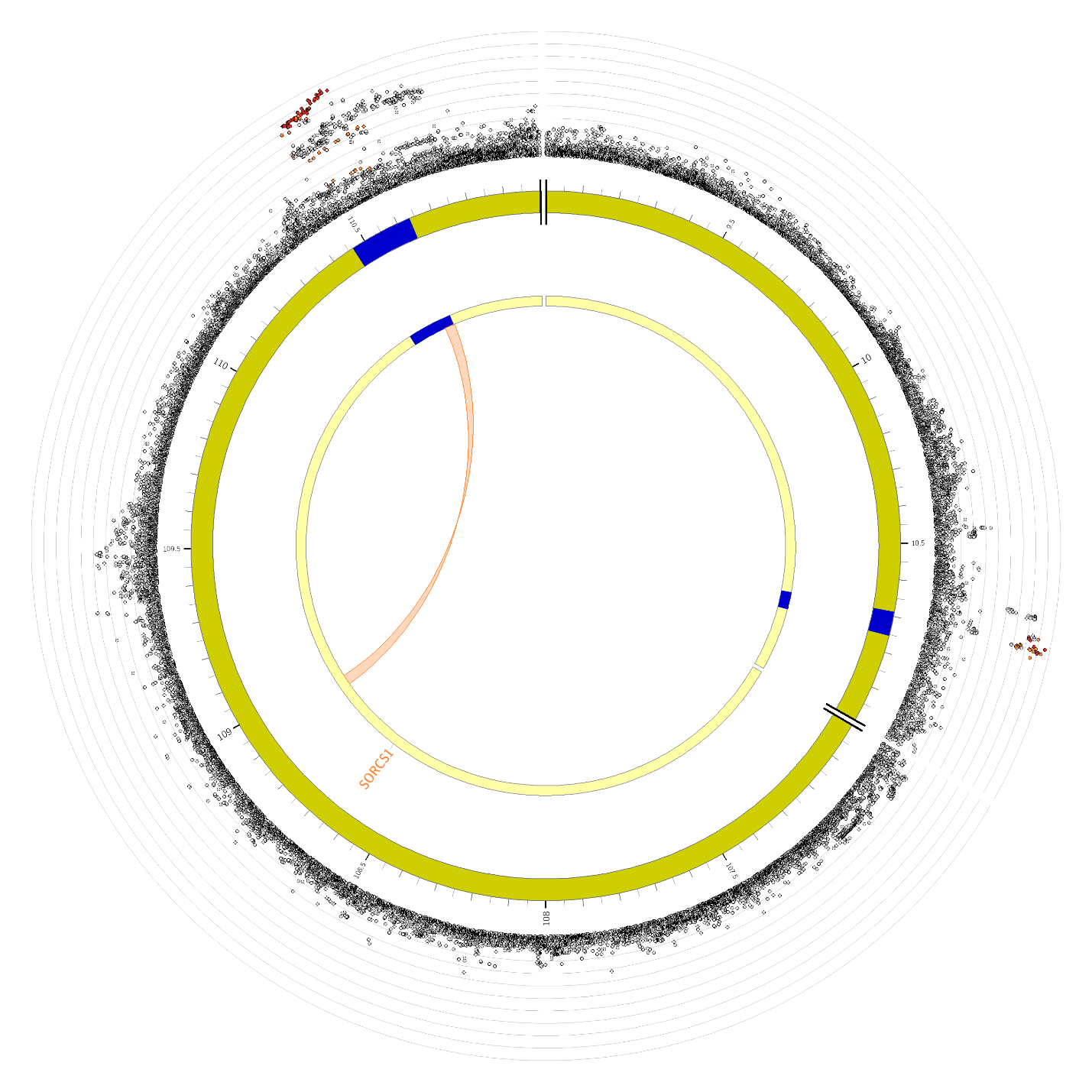

(g) **Chromosome 11**

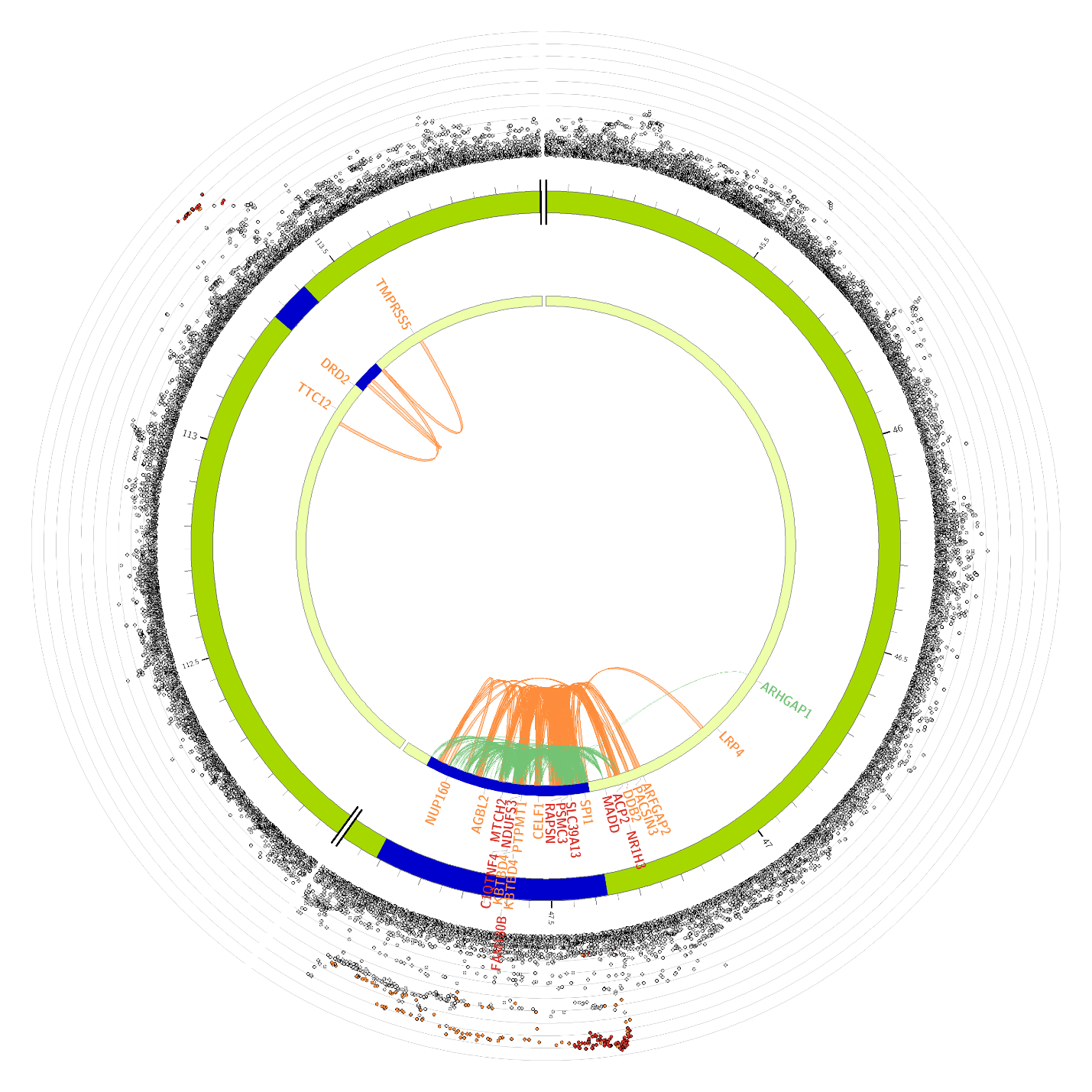

(h) **Chromosome 17**

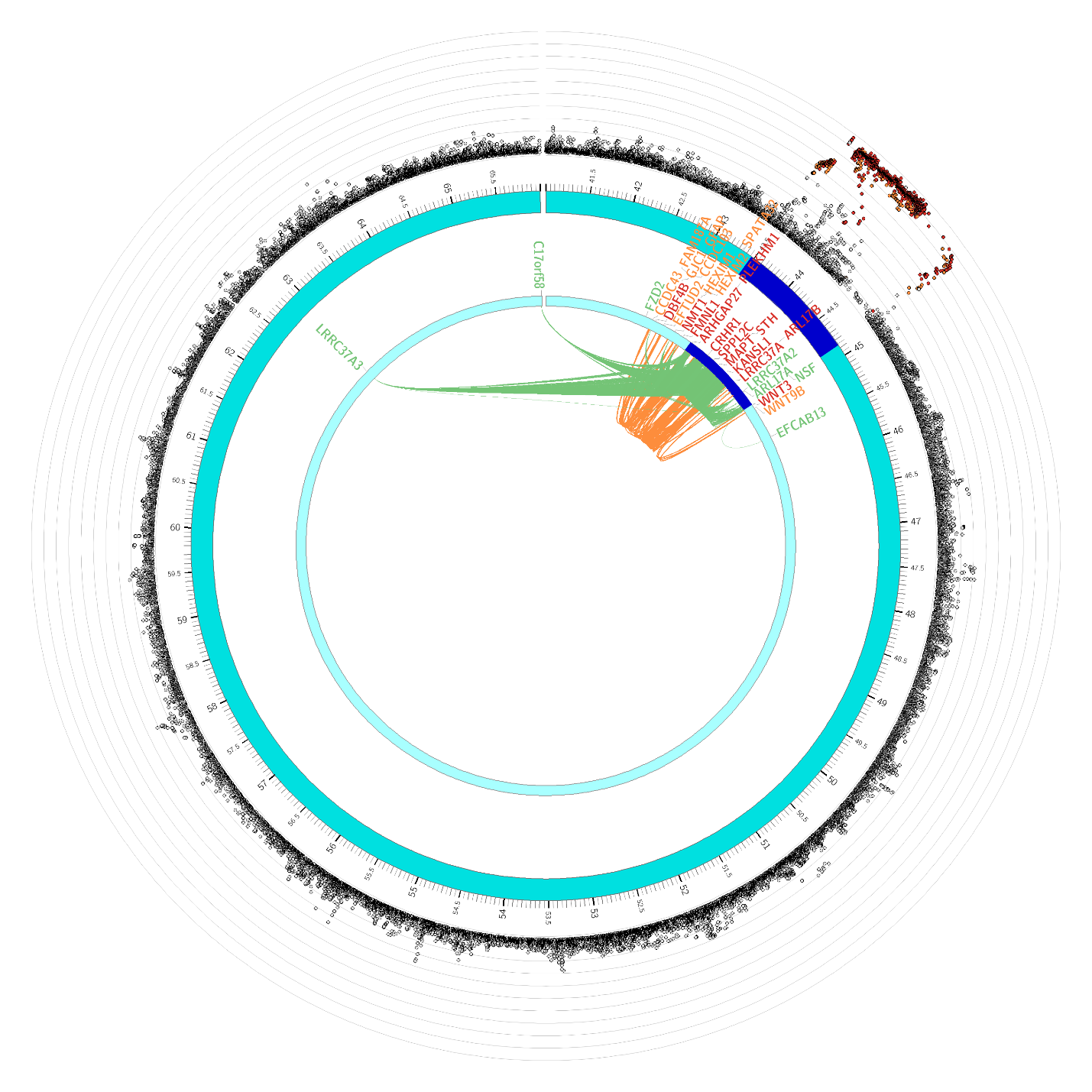

(i) **Chromosome 19**

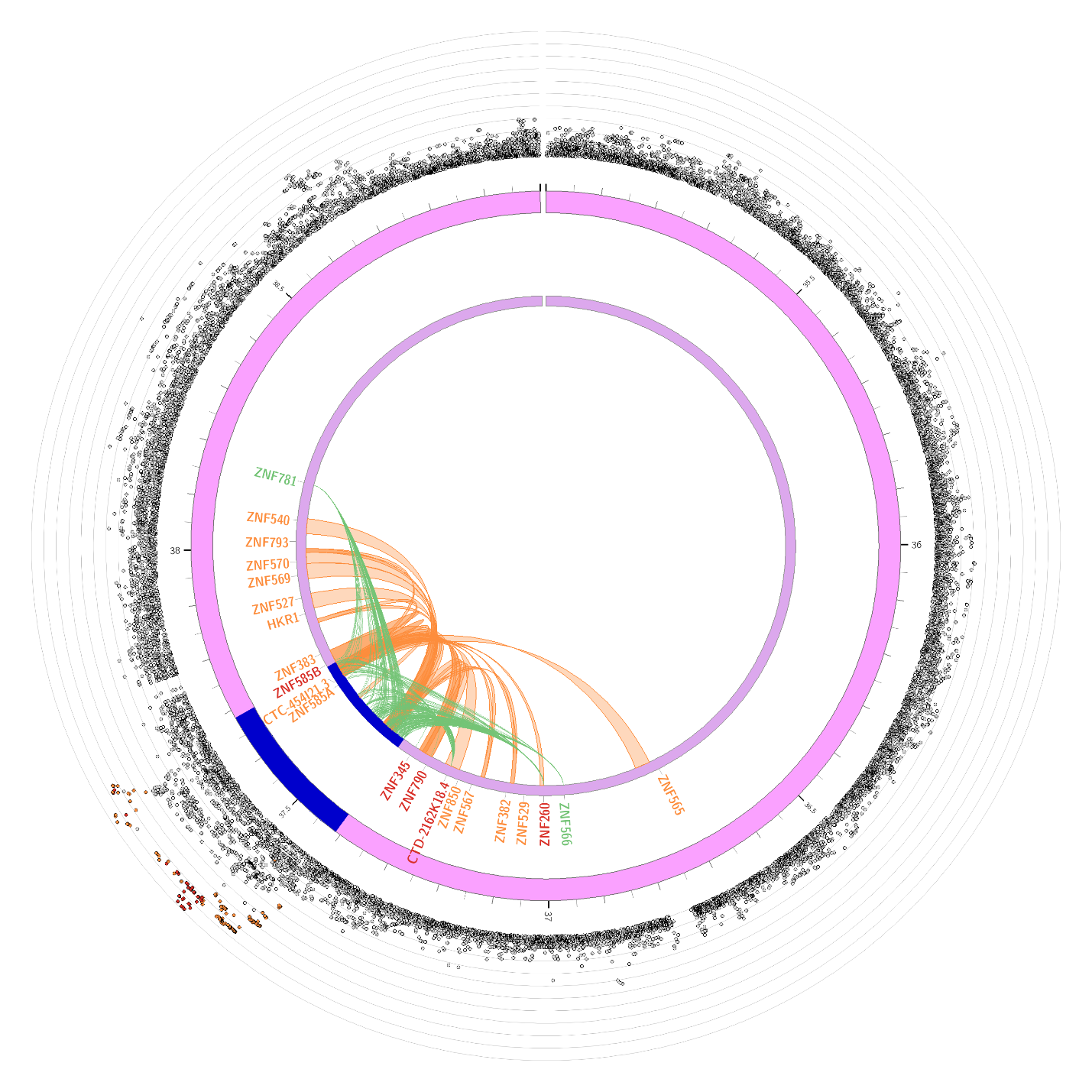

**Supplemental Figure 19: Circos plots for chromosomes containing genome-wide significant loci for the MaxAlc MTAG GWAS results (MaxAlc-PAU).**

**Note. Outer most layer** is a Manhattan plot of genome-wide association study (GWAS) single-nucleotide polymorphisms (SNPs) with *p*≤0.05. SNPs are plotted by chromosomal position along the *x*-axis with their corresponding -log^-10^ *p*-value on the *y*-axis. Linkage-disequilibrium (LD) between the identified lead SNP and surrounding SNPs is indicated from r^2^>0.8 (red), r^2^>0.6 (orange), r^2^>0.4 (green), r^2^>0.2 (blue). SNPs that are not in LD with the lead SNP (r^2^≤0.02) are gray. **Second layer (chromosome ring):** Chromosomal regions containing identified genomic risk loci are colored in blue. The names of genes implicated based upon variant associations with brain tissue expression quantitative trait loci (eQTLs) are colored green. The names of genes implicated based upon 3D chromatin interactions (Hi-C) are colored orange. Genes that are mapped based upon both eQTLs and Hi-C associations are colored red. **Third layer (chromosome ring):** Variants mapped to genes based upon associations with brain tissue eQTLs are linked in green. Variants mapped to genes based upon Hi-C data are linked in orange.

(a) **Chromosome 1**

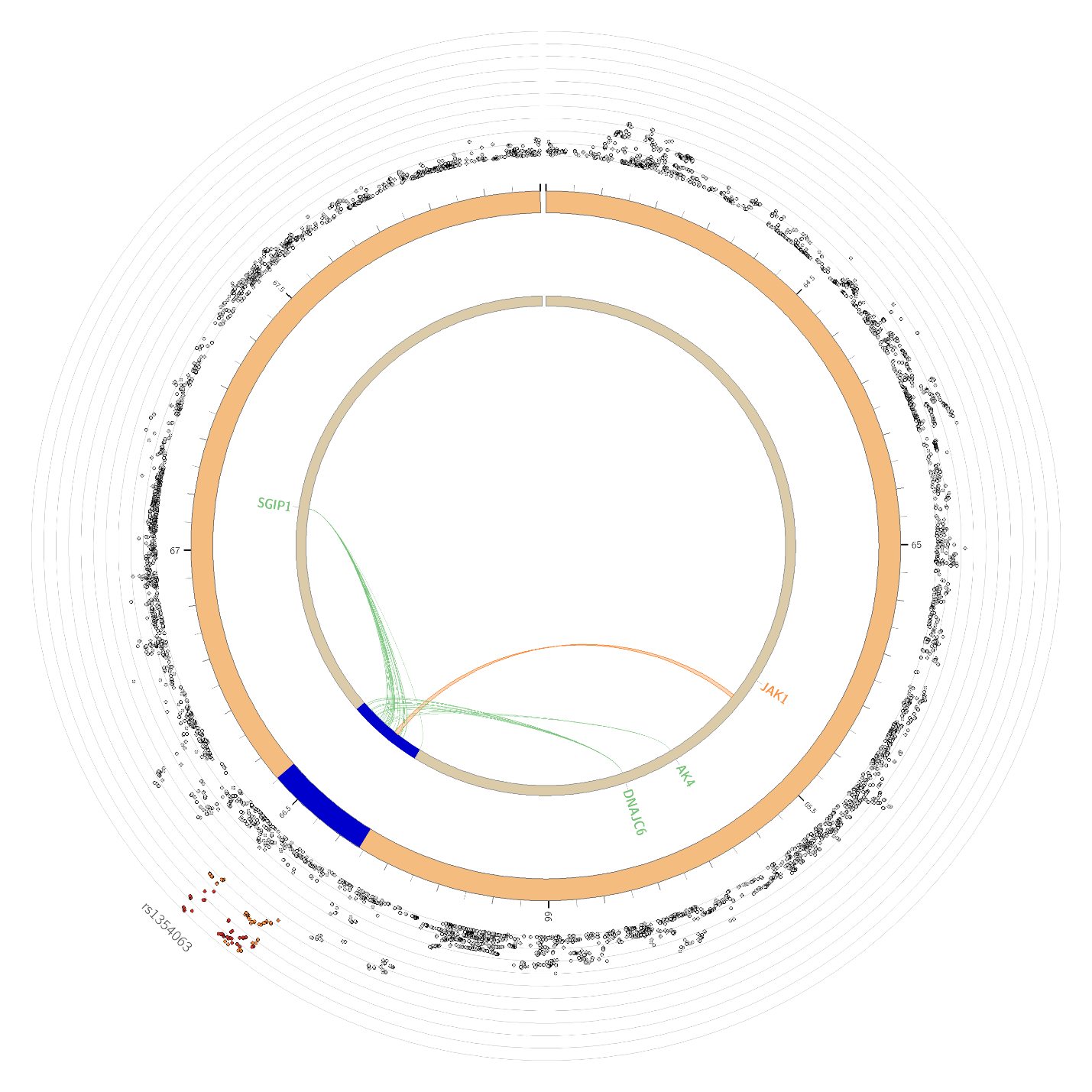

(b) **Chromosome 2**

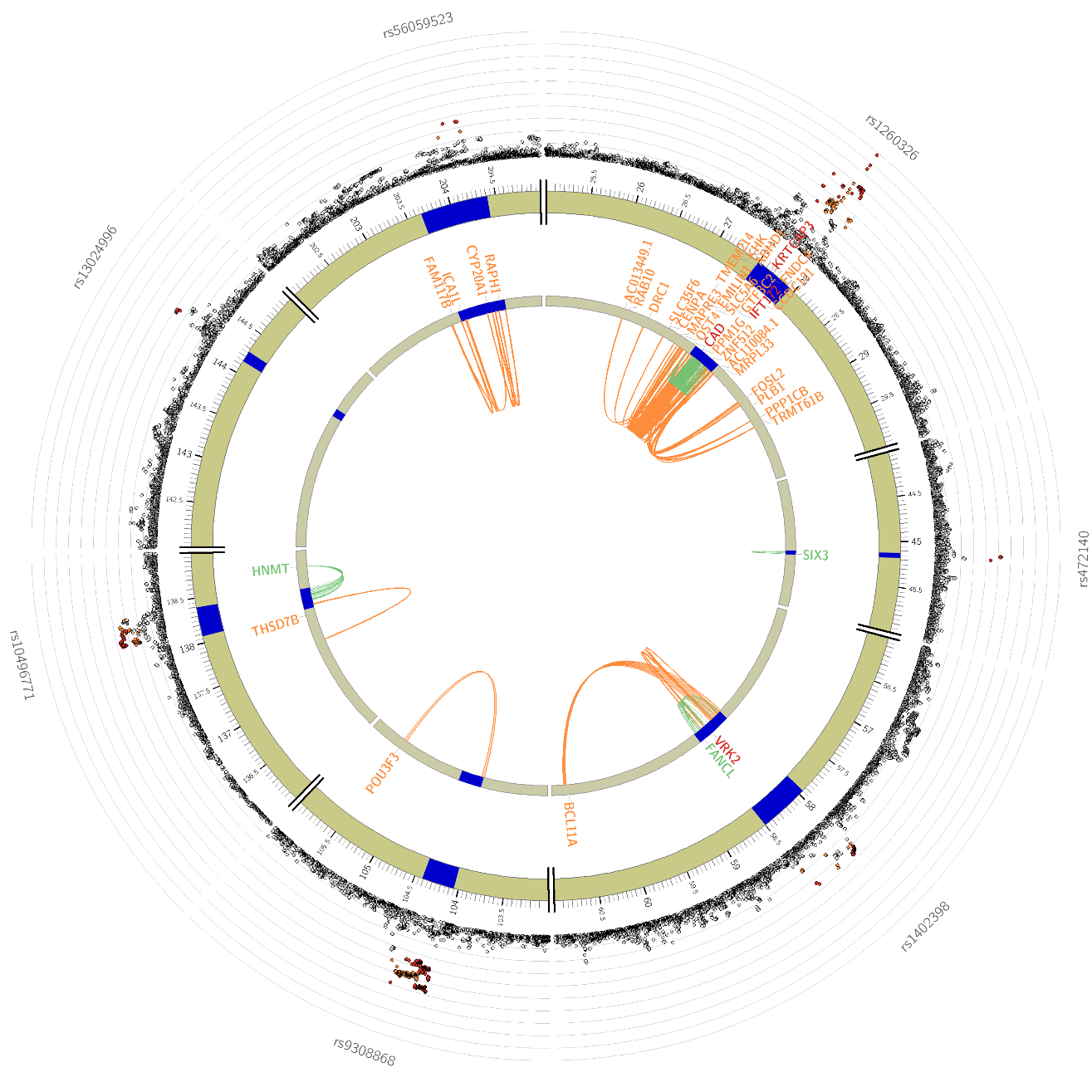

(c) **Chromosome 3**

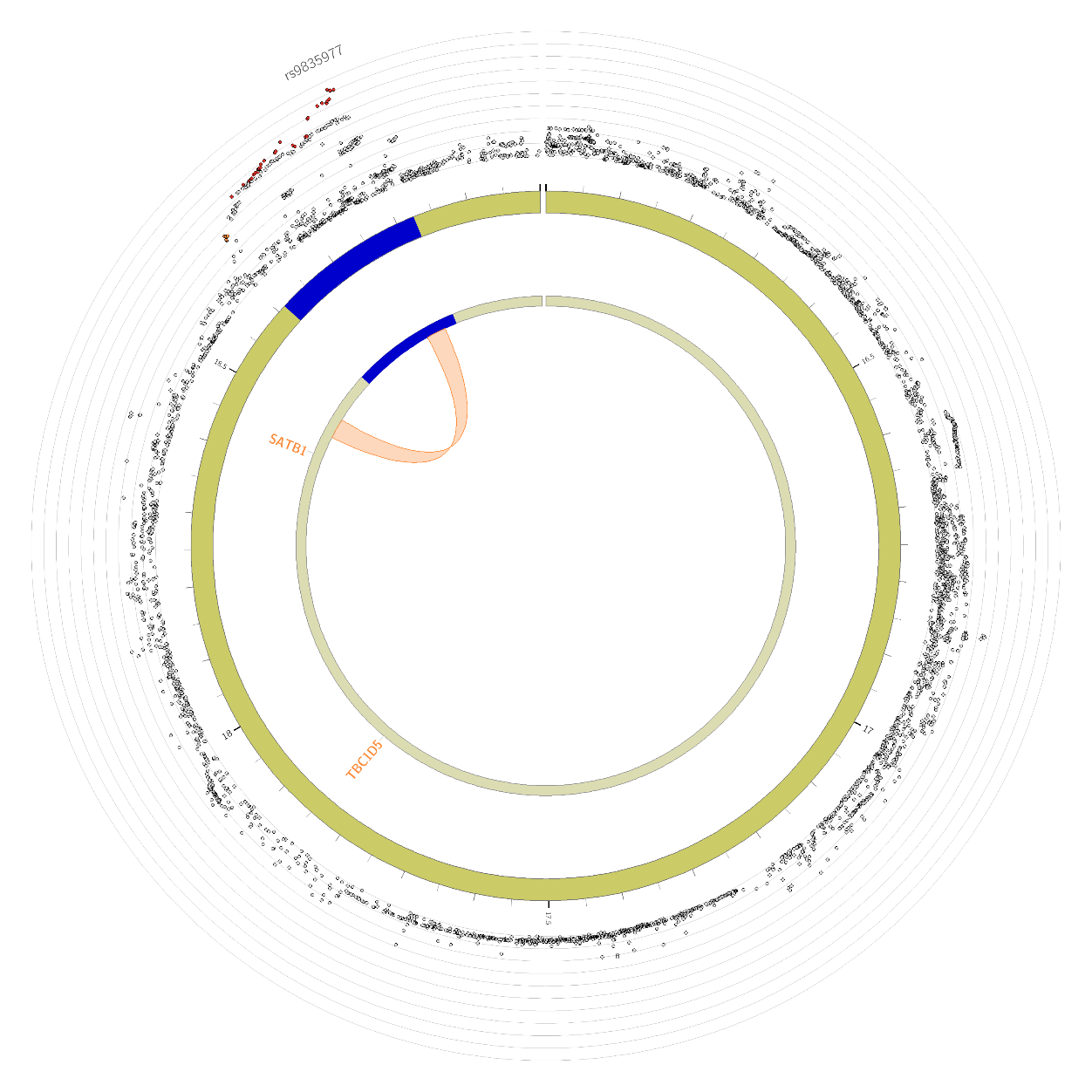

(d) **Chromosome 4**

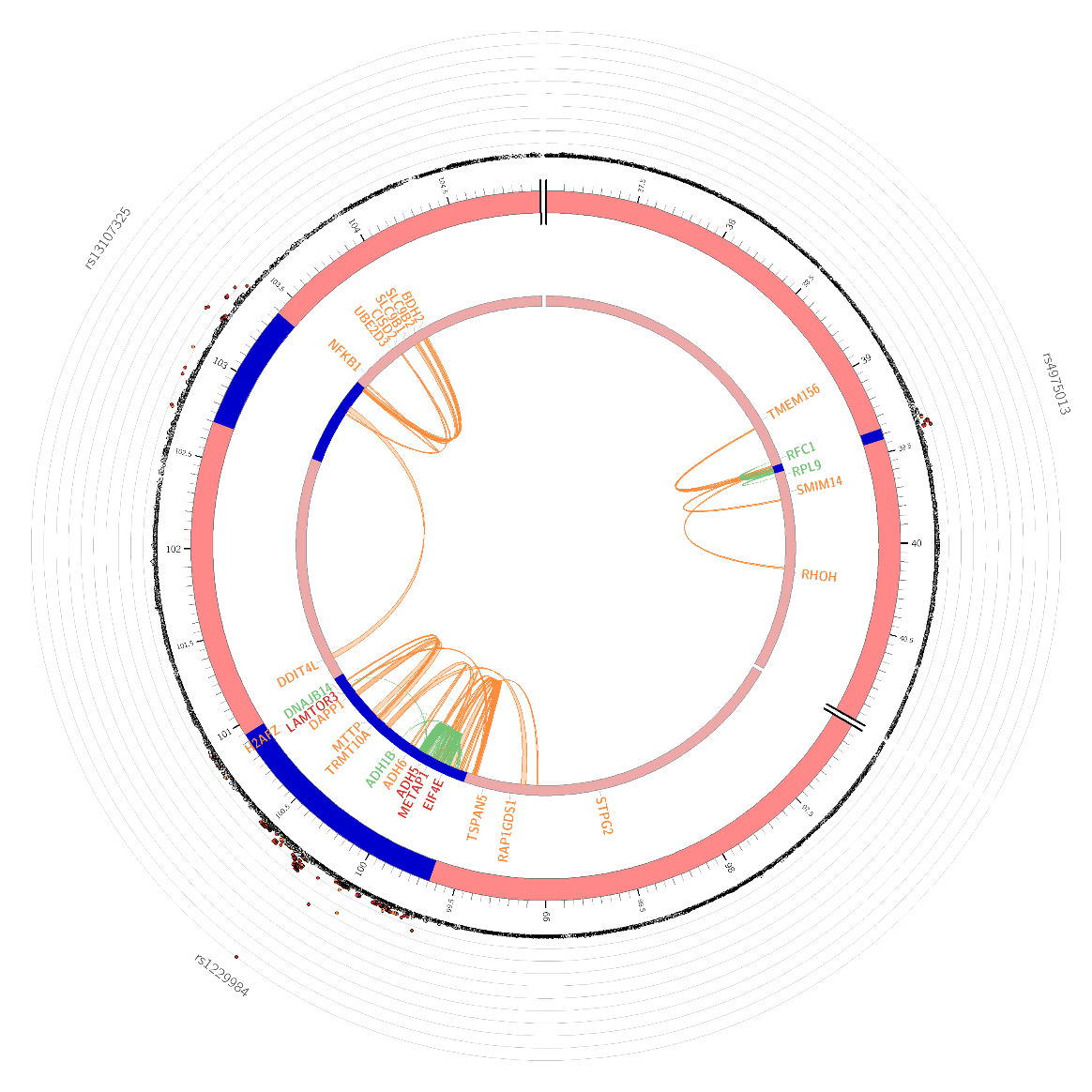

(e) **Chromosome 5**

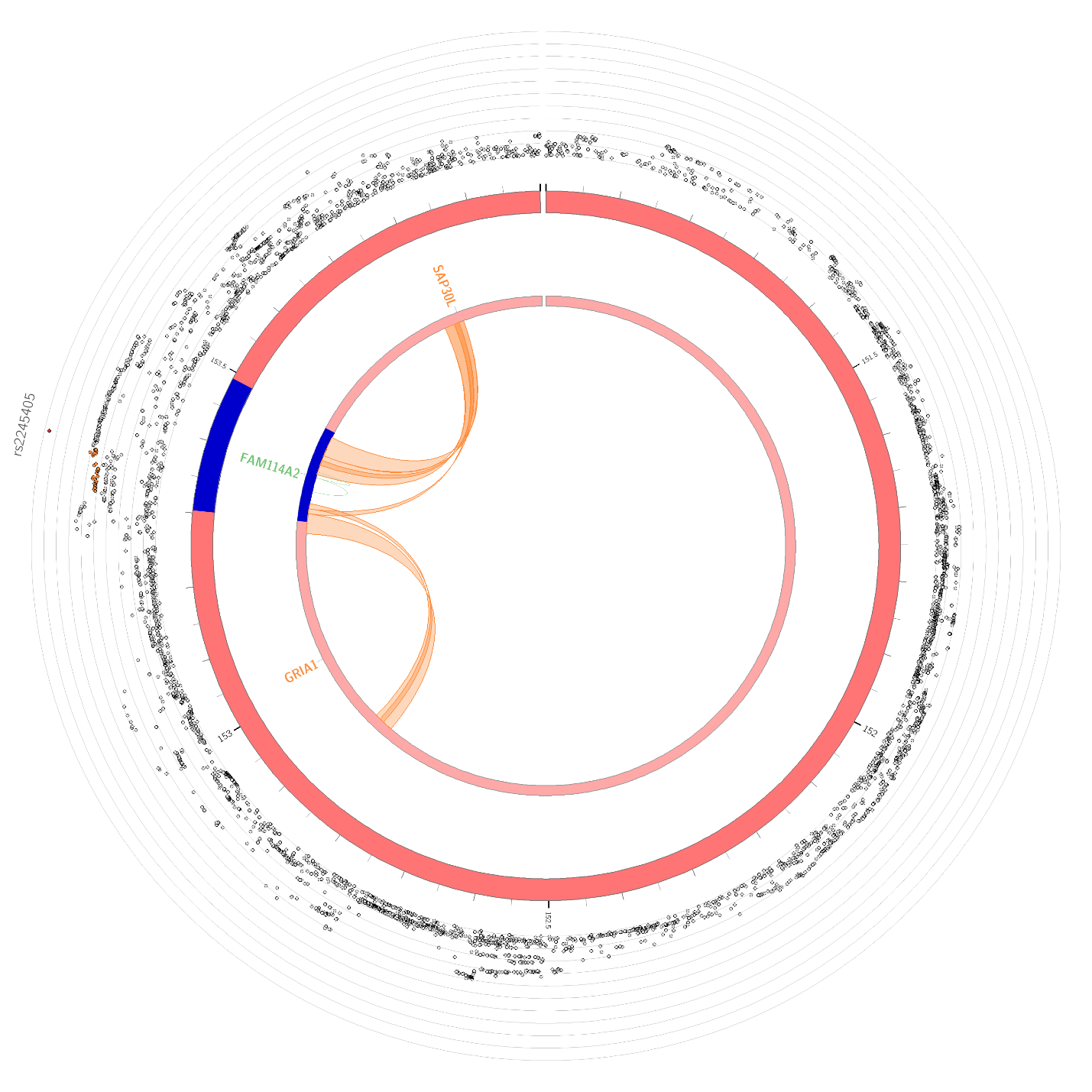

(f) **Chromosome 7**

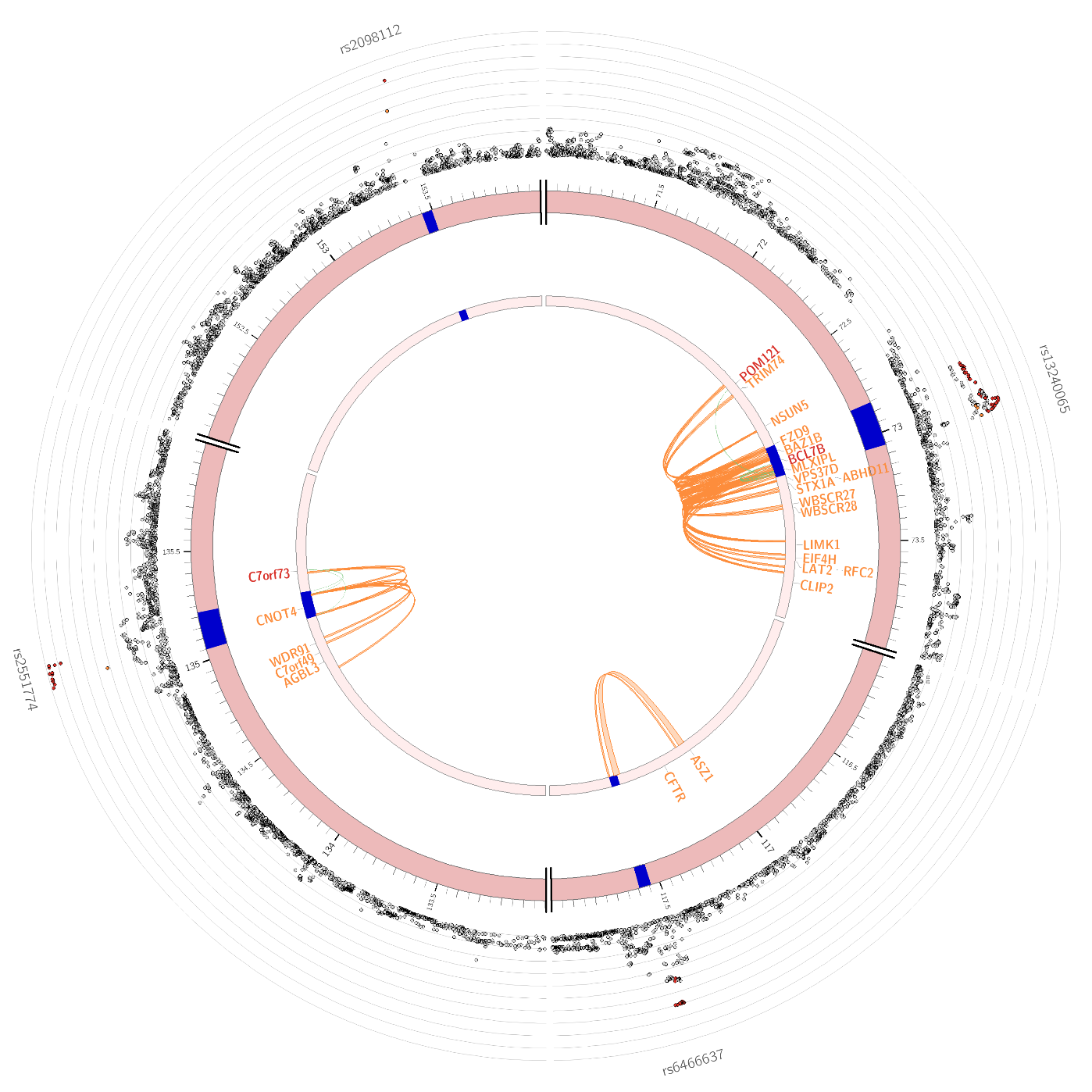

(g) **Chromosome 8**

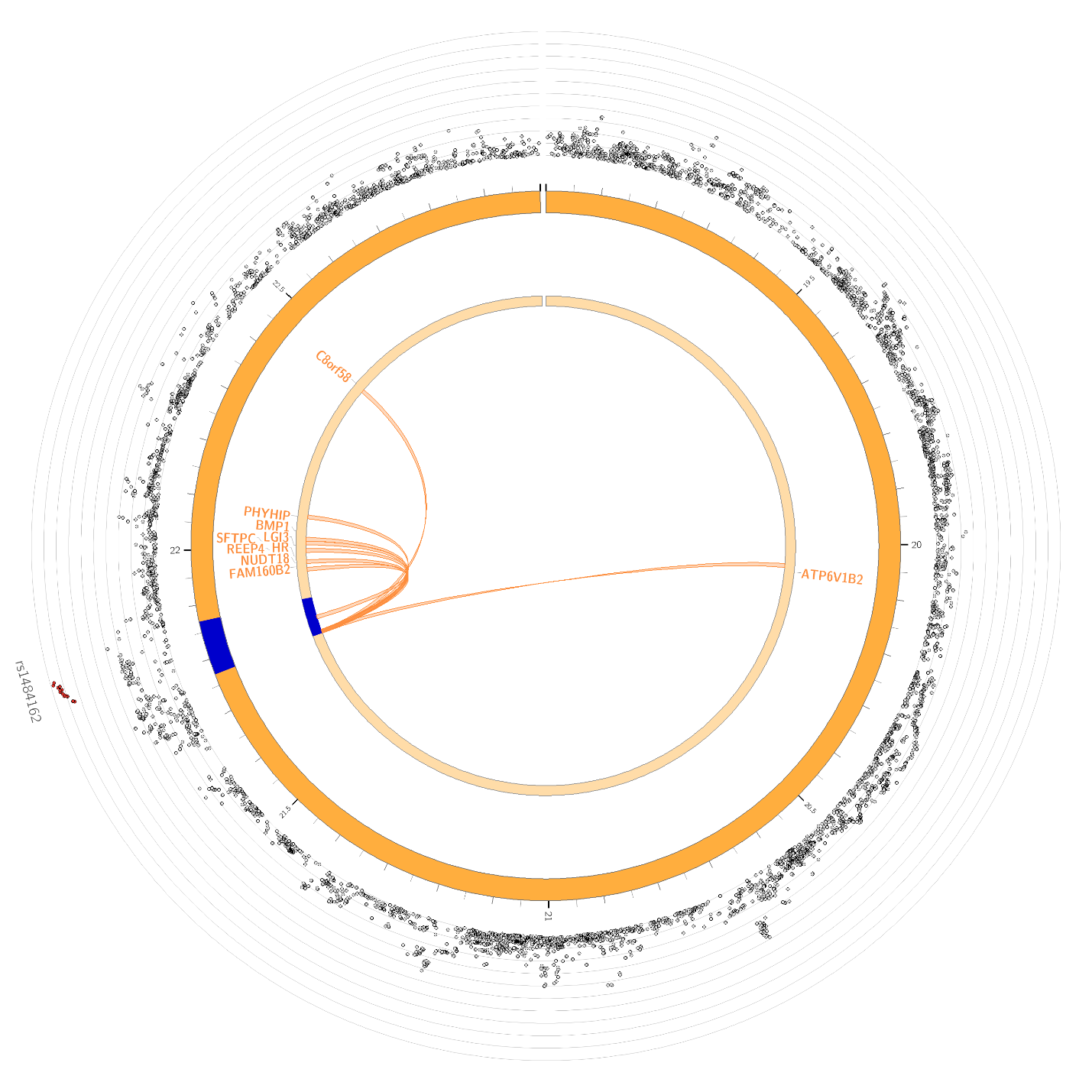

(h) **Chromosome 10**

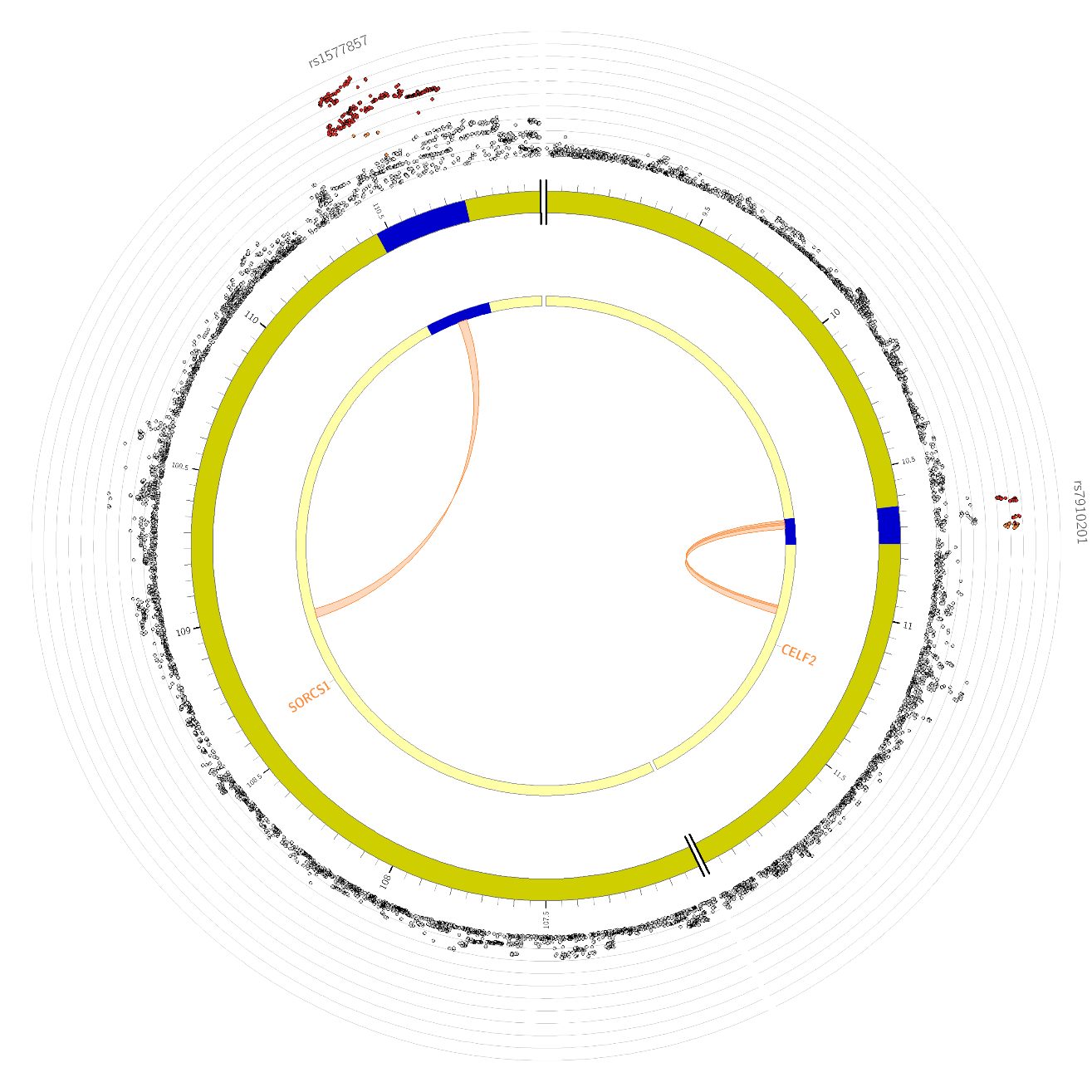

(i) **Chromosome 11**

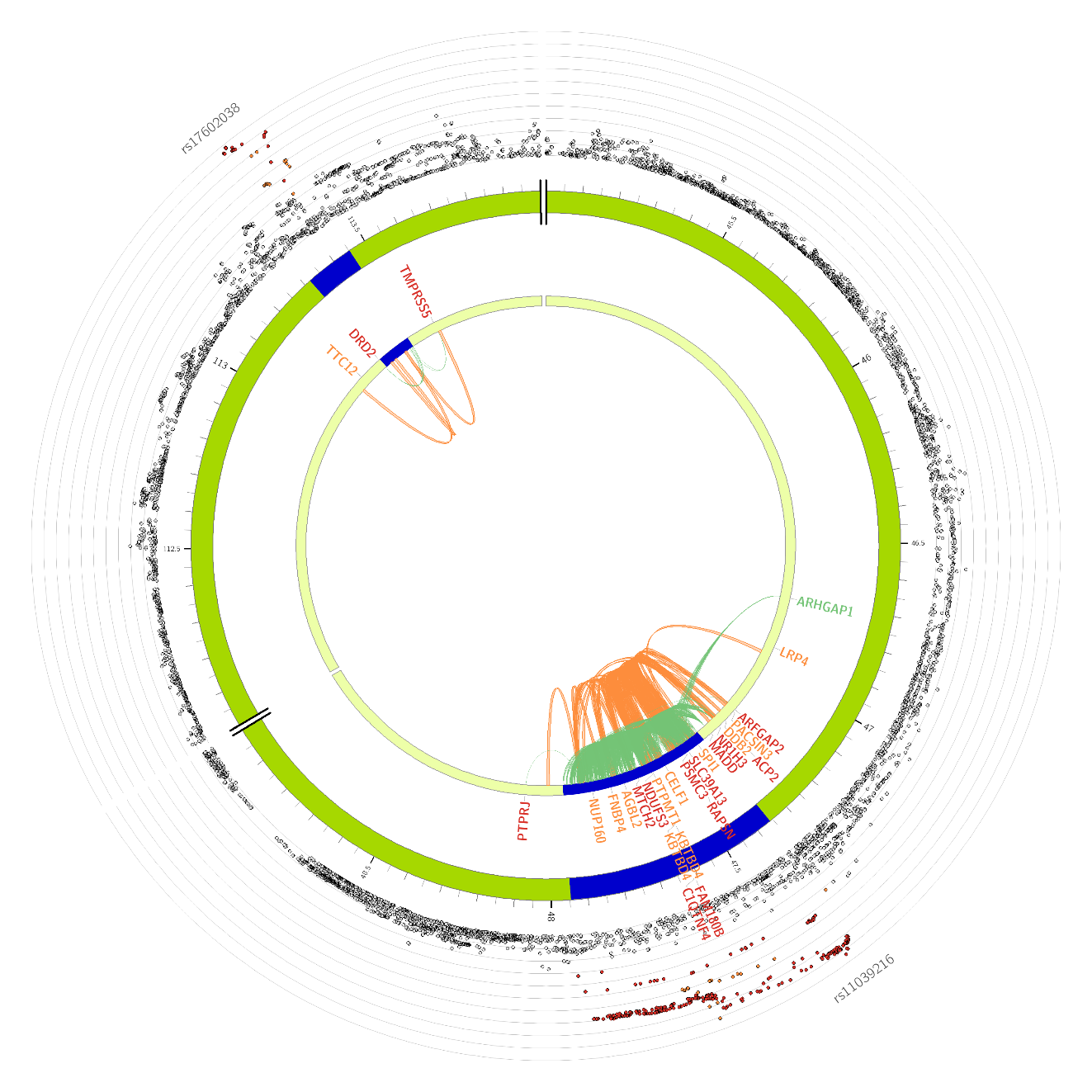

(j) **Chromosome 12**

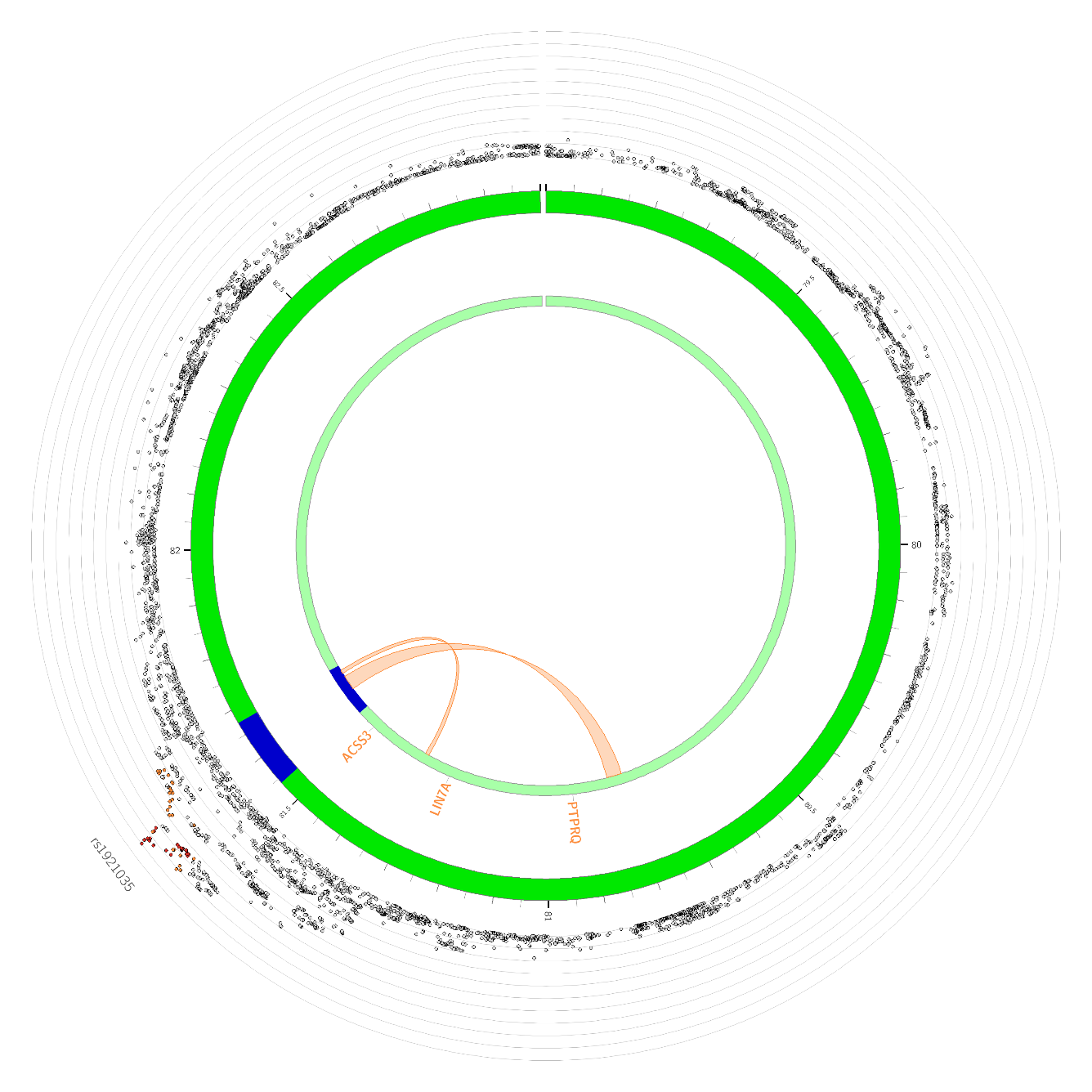

(k) **Chromosome 13**

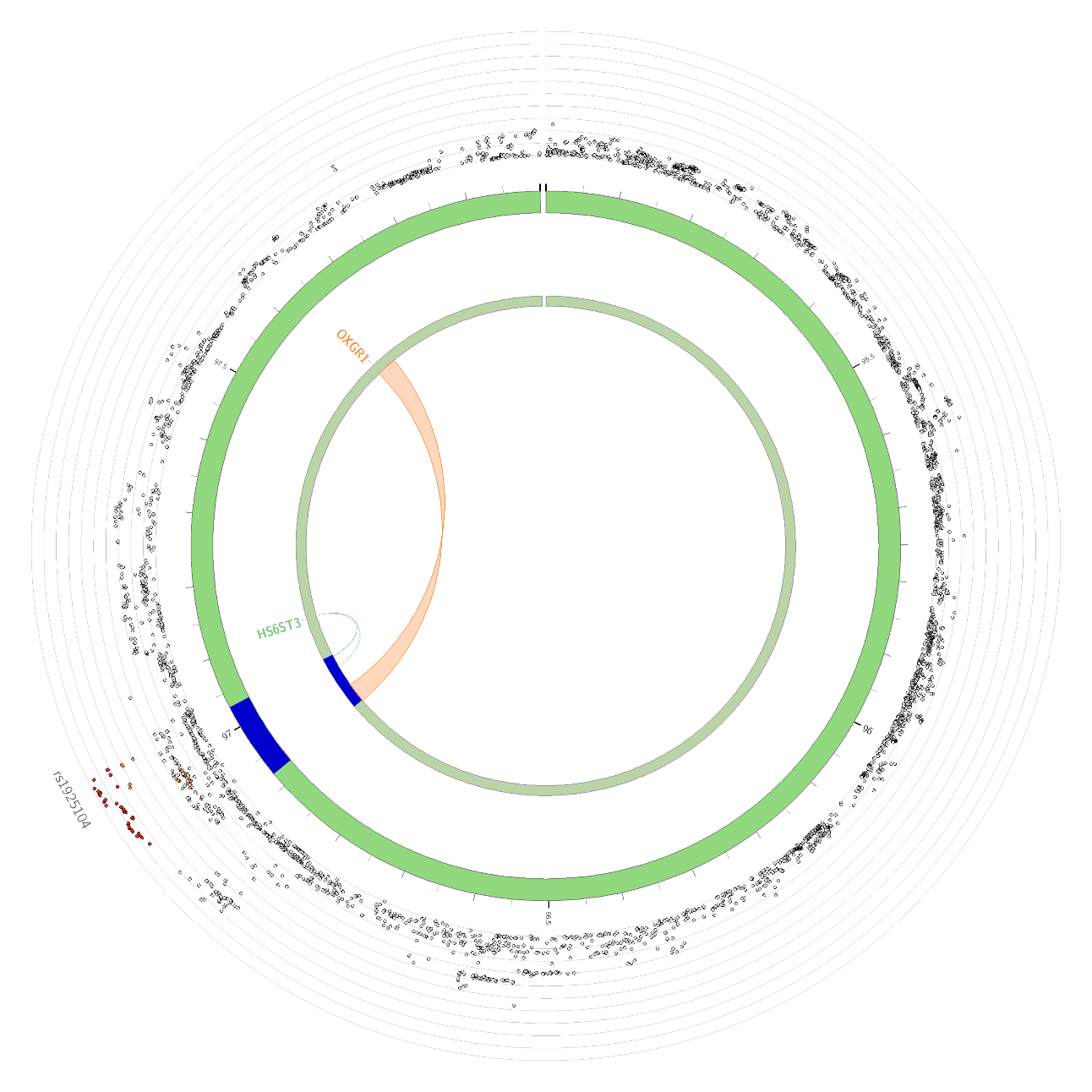

(l) **Chromosome 14**

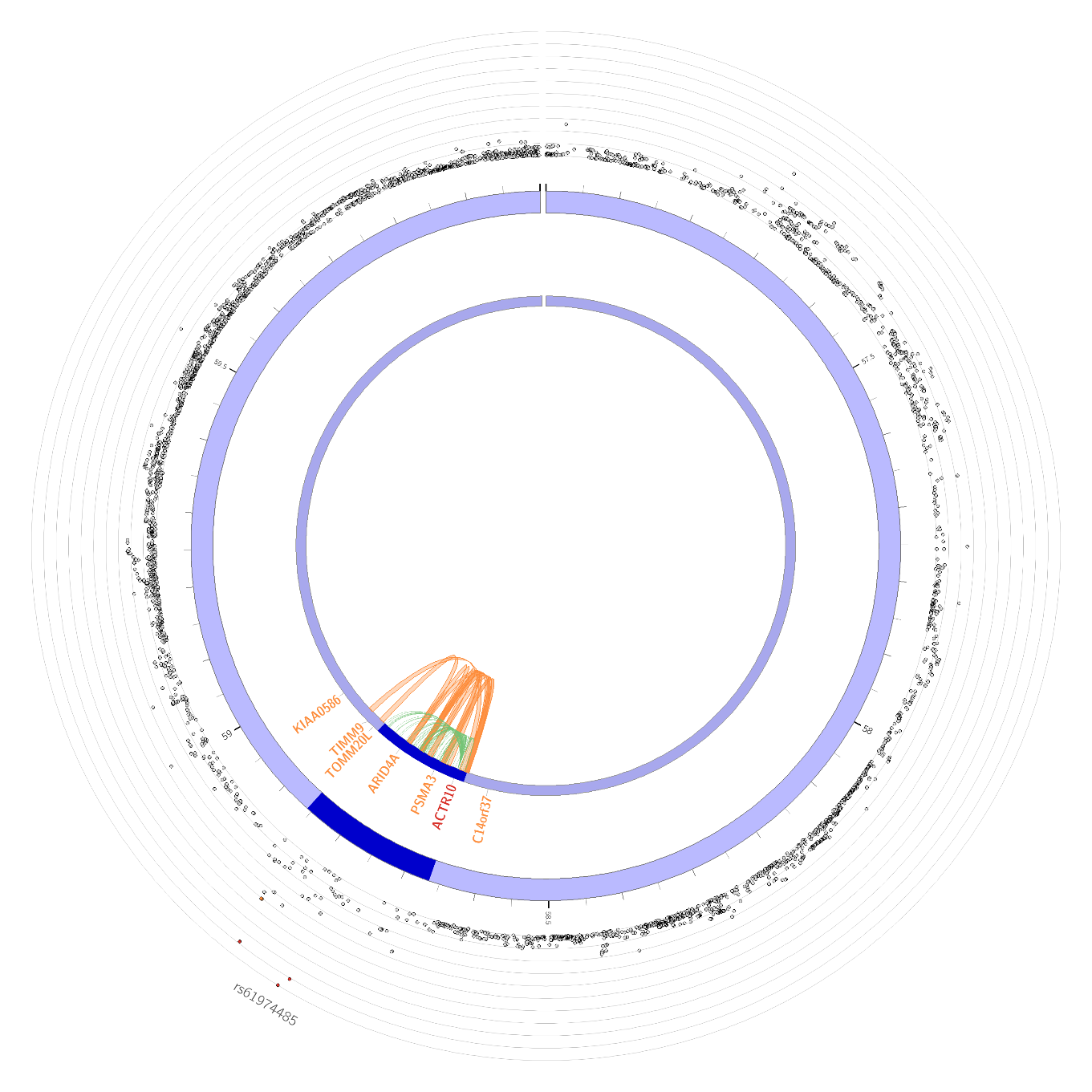

(m) **Chromosome 15**

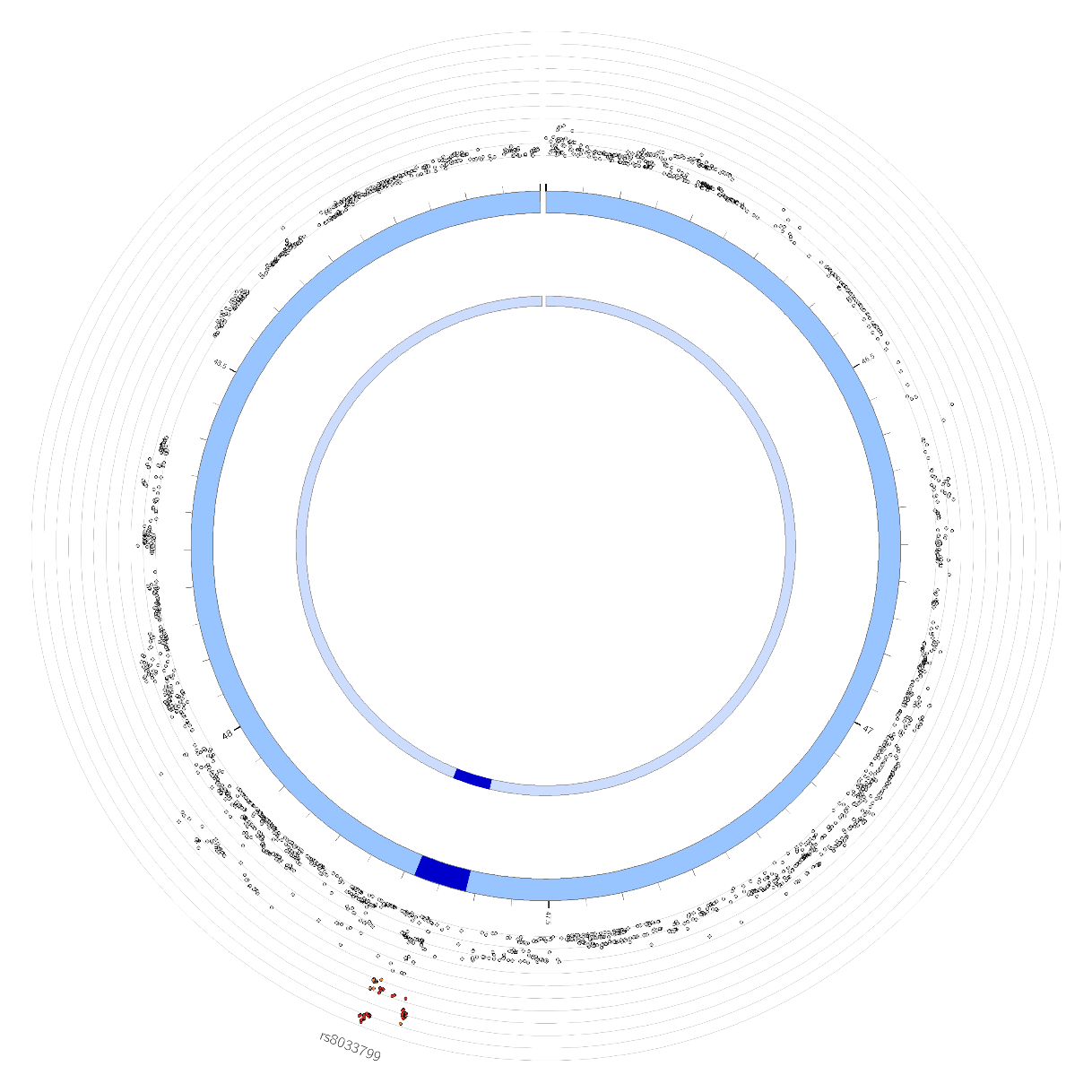

(n) **Chromosome 16**

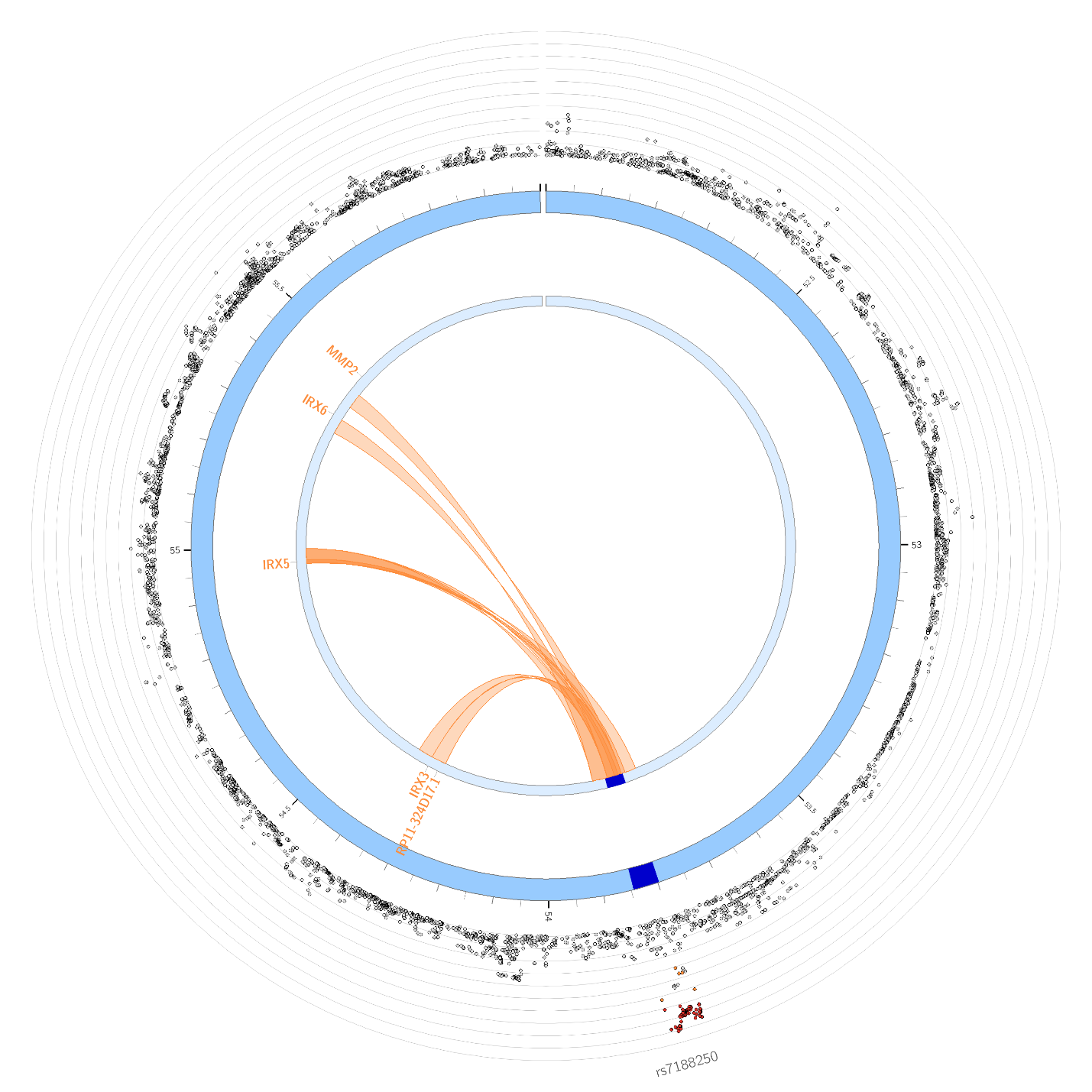

(o) **Chromosome 17**

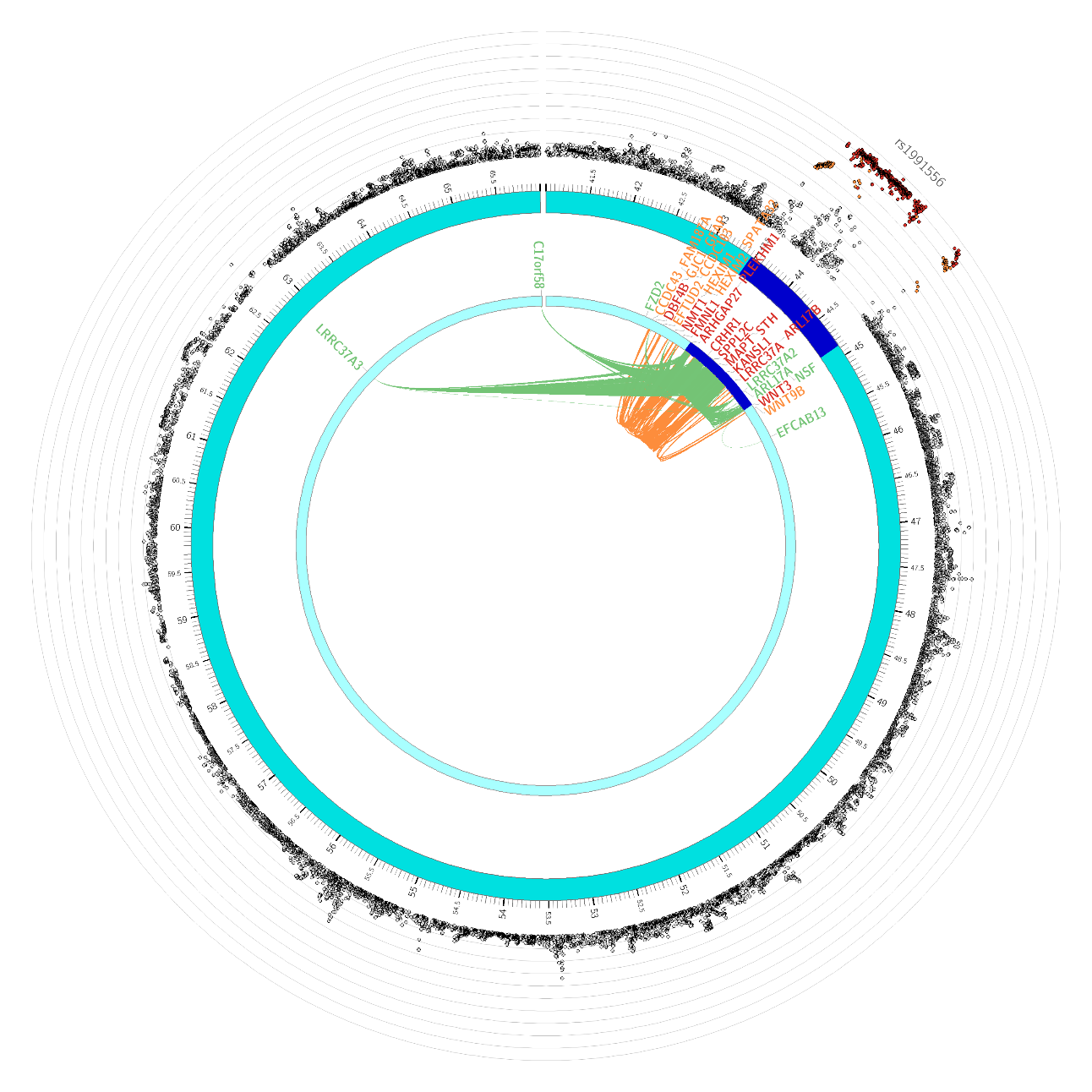

(p) **Chromosome 19**

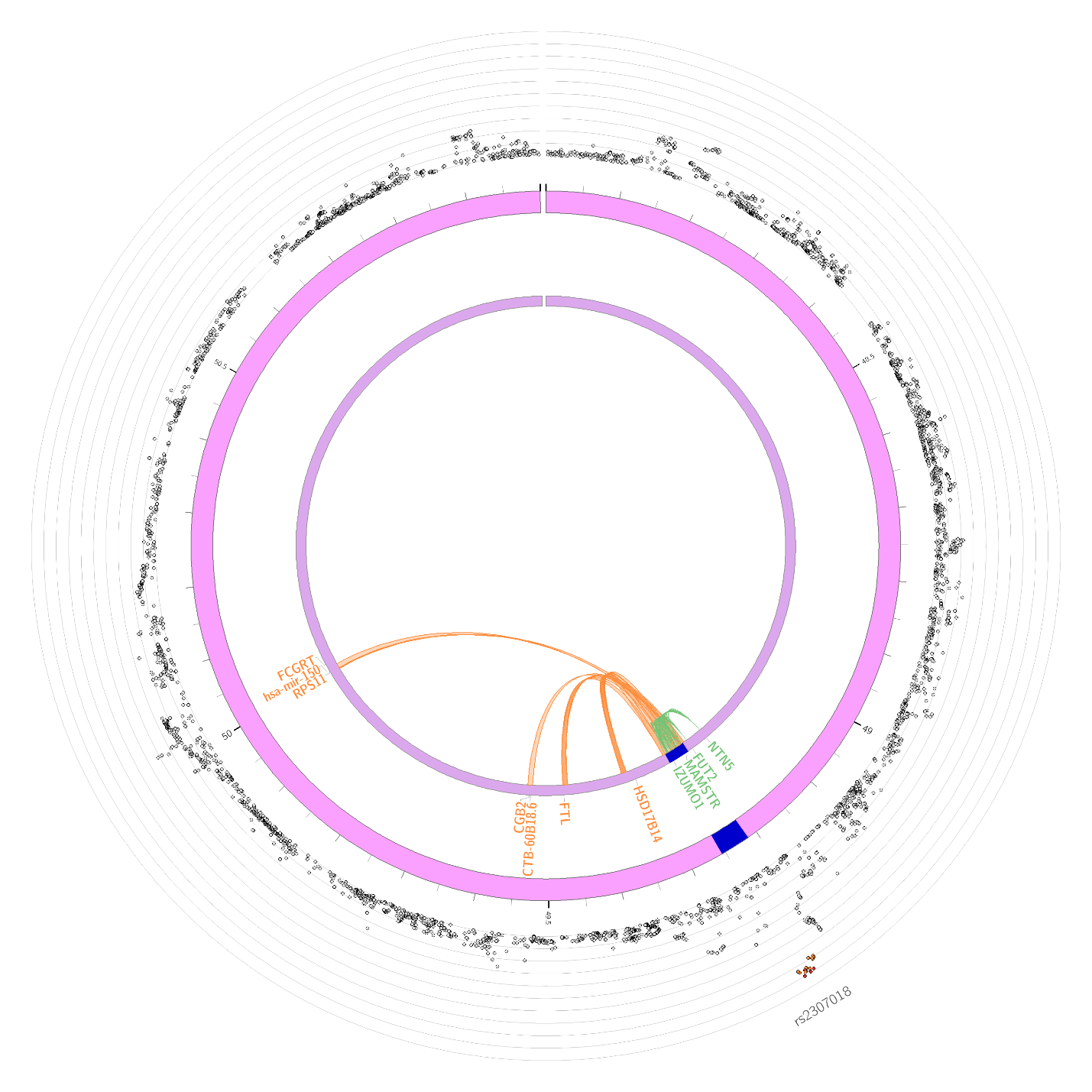

(q) **Chromosome 20**

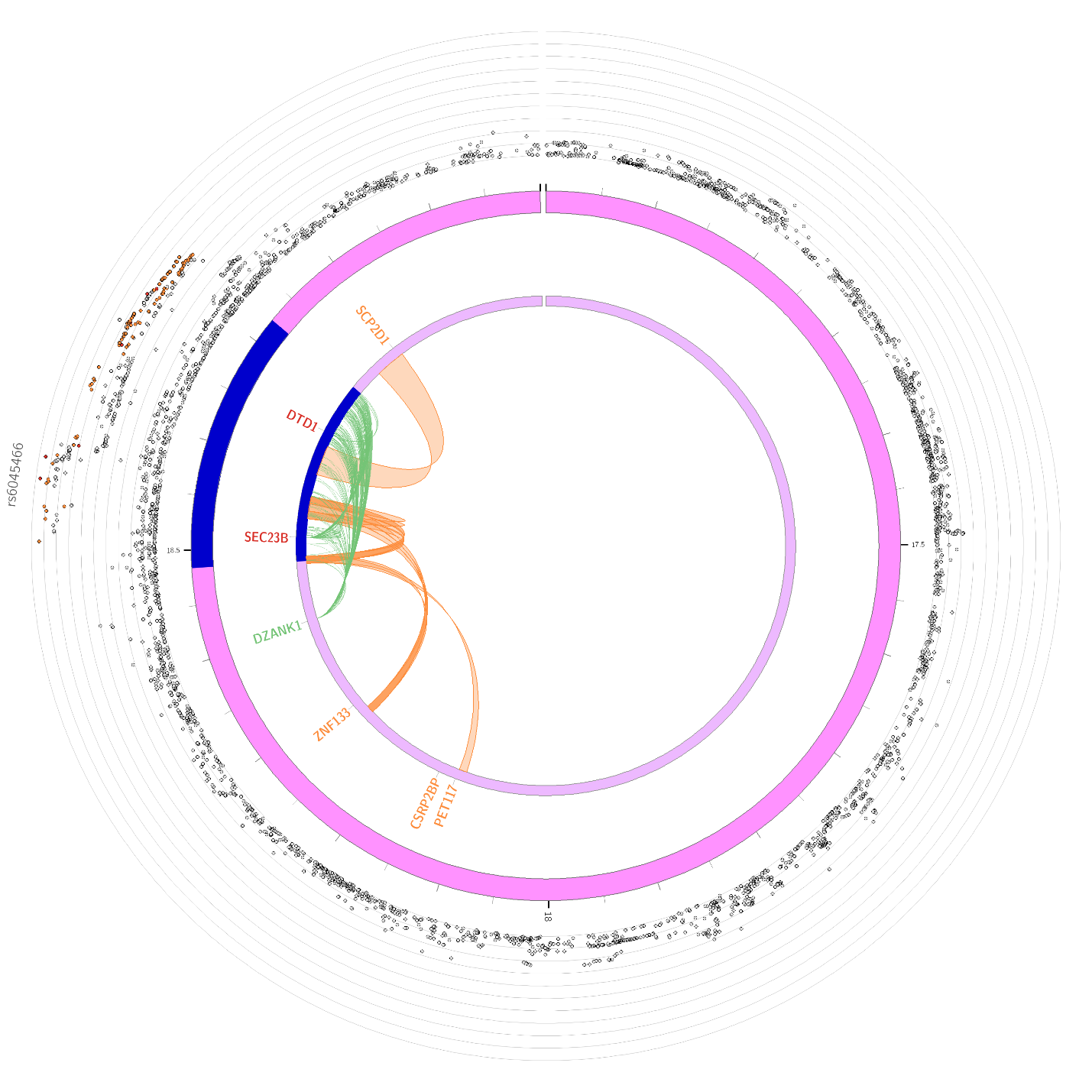

(r) **Chromosome 22**

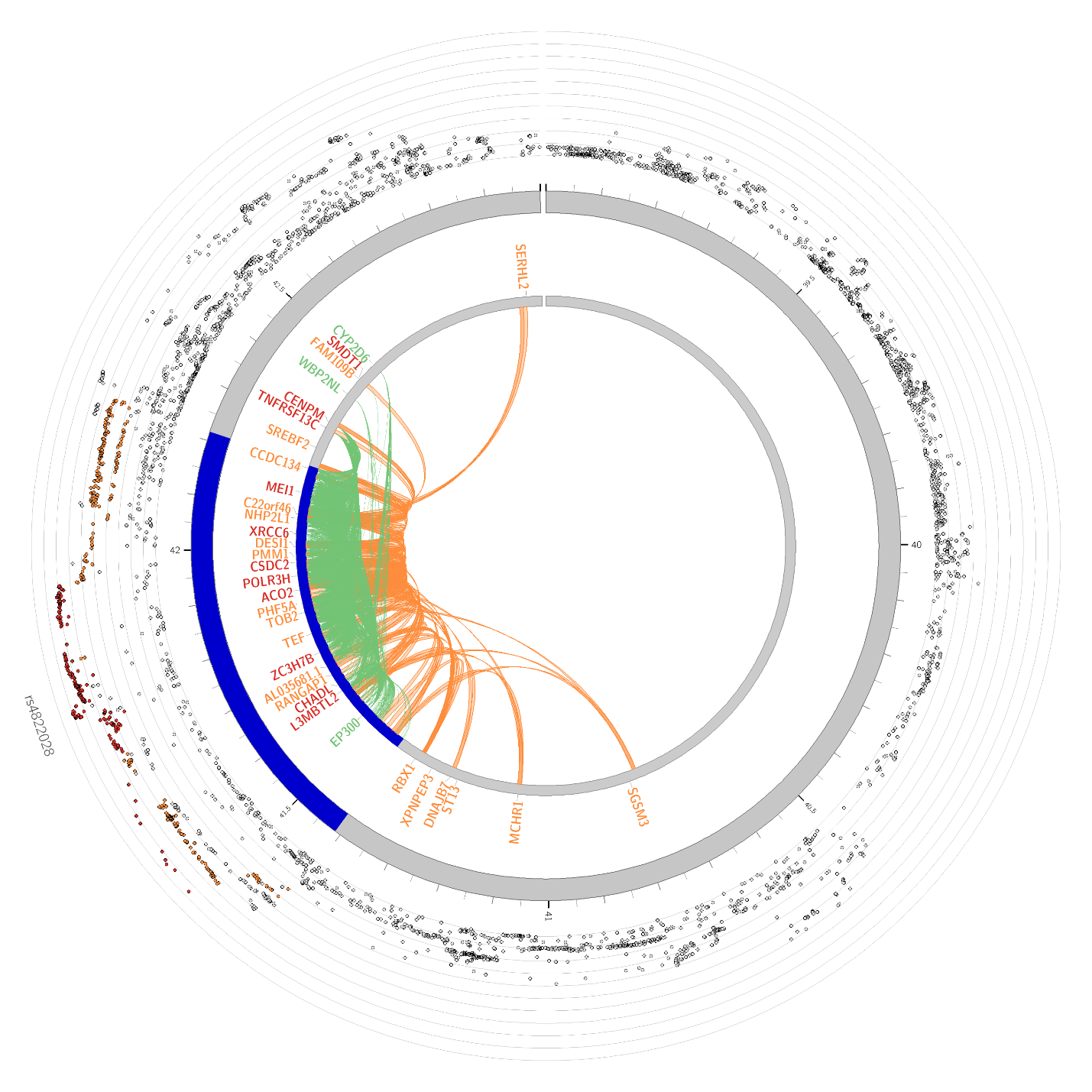
